# Changes in health behaviors, mental and physical health among older adults under severe lockdown restrictions during the COVID-19 pandemic in Spain

**DOI:** 10.1101/2021.02.15.21251738

**Authors:** Esther García-Esquinas, Rosario Ortolá, Iago Gine-Vázquez, José A Carnicero, Asier Mañas, Elvira Lara, Alejandro Alvarez-Bustos, German Vicente-Rodriguez, Mercedes Sotos-Prieto, Beatriz Olaya, Francisco José Garcia-Garcia, Narcis Gusi, Jose R. Banegas, Irene Rodríguez-Gómez, Ellen A. Struijk, David Martínez-Gómez, Alberto Lana, Josep María Haro, José L. Ayuso-Mateos, Leocadio Rodríguez-Mañas, Ignacio Ara, Marta Miret, F Rodríguez-Artalejo

## Abstract

**Background:** We aimed to examine main changes in health behaviors, mental and physical health among older adults under severe lockdown restrictions during the COVID-19.

**Methods:** We used prospective data from 3041 participants in four cohorts of community-dwelling individuals aged ≥65 years in Spain. Data were obtained using validated questionnaires through a pre-pandemic face-to-face interview and a telephone interview conducted between weeks 7 to 15 after the beginning the COVID-19 lockdown. Lineal or multinomial, as appropriate, regression models with adjustment for the main confounders were used to assess changes in the outcome variables from the pre-pandemic to the confinement period, and to identify their associated factors.

**Results:** On average, the confinement was not associated with a deterioration in lifestyle risk factors (smoking, alcohol intake, diet or weight), except for a decreased physical activity and increased sedentary time, which reversed with the end of confinement. However, chronic pain worsened, and moderate declines in mental health, that did not seem to reverse after restrictions were lifted, were observed. Several subgroups of individuals were at increased risk of developing unhealthier lifestyles or mental health decline with confinement: (i)-males (for physical activity and sedentariness), (ii)-those with greater social isolation (for diet, physical activity, mental health), (iii)-feelings of loneliness (for diet, sleep quality, mental health), (iv)-poor housing conditions (for diet, physical activity, TV viewing time), (v)-unhealthy sleep duration (for physical activity and sedentariness), and (vi-worse overall health or chronic morbidities (for physical activity, screen time, mental health). On the other hand, previously having a greater adherence to the Mediterranean diet and doing more physical activity protected older adults from developing unhealthier lifestyles with confinement.

**Conclusions:** The lockdown during the first wave of the COVID-19 in Spain, which was one of the most restrictive in Europe, only led to minor average changes in health behaviors among older adults. However, mental health was moderately affected. If another lockdown were imposed on this or future pandemics, public health programs should specially address the needs of older individuals with male sex, greater social isolation, poor housing conditions and chronic morbidities, because of their greater vulnerability to the enacted movement restrictions

## INTRODUCTION

The COVID-19 pandemic has rocked our society to its core. Until a safe and effective vaccines are globally distributed, social distancing interventions to reduce the transmission of SARS-CoV-2 are essential (1), but themselves are not extent from health and economic challenges (2–5).

The State of Alarm imposed by the Spanish Government to curb the spread of the SARS-CoV-2 infection, entailed a national lockdown starting on March 15^th^, and the imposition of distancing measures like the cancelation of mass events or the closure of non-essential customer-facing business and educational institutions (6).Two weeks later, on March 29^th^, the Government strengthened these measures to avoid the saturation of intensive care units (7). A period of five weeks started in which citizens were only allowed to leave their homes for essential work, buying of staple products or emergencies. On May 4^th^, citizens were first authorized to leave their homes to exercise or walk, for a maximum of 1 hour a day, following timetables according to age, and only being accompanied by people they had been living together during the lockdown. From May 10^th^ to June 21^st^, a progressive de-escalation of confinement measures lead to the so-called “New Normality” in which Spaniards were allowed to attend their jobs, gather in small groups, and move between provinces as long as they complied with safe distancing and face covering requirements. All around the world, countries dealt with the first wave of the pandemic with different degree of movement restrictions, but the Spanish lockdown, especially during those five weeks between March 30^th^ and May 4^th^, was one of the most restrictive in Europe.

A few studies have investigated the health consequences of the lockdown during the first wave of the pandemic in the Spanish adult population. Using convenience sample surveys conducted between March and June 2020, in which participants reported their perceived changes in lifestyle and health-related factors since the start of the pandemic, these studies have shown a general worsening in psychological and mental health (8–10), as well as in sleep quality (11–13), mild or no changes in tobacco and alcohol consumption (11,14), slight or no changes in weight (14–16), conflicting results for diet quality (17,18), important reductions in physical activity and increases in sedentary time (11,19–21). Interestingly, one survey found that the percentage of interviewees meeting the guidelines regarding screen time became lower between March 22^nd^ and April 5^th^, while unhealthy alcohol consumption and insufficient physical activity decreased during this three-week period (22).

Unfortunately, none of these studies described the determinants of the observed changes in health behaviors, most were at high risk of information bias (i.e. people not accurately reporting their past exposures or symptoms, especially at a time of emotional distress), and, although a few provided stratified information by age groups, most presented aggregated information for young, middle-aged and older adults. Moreover, most of these studies were based on convenience samples, which increase the risk of selection bias. As the COVID-19 epidemic evolves and new social distancing measures may be needed (23), and being aware of the risk of future infectious pandemics, it is critical that we identify population subgroups that are at increased risk of health deterioration with social isolation and confinement. This is especially important for older adults, because they represent around 10% of the worldwide population (up to 20% in Spain), frequently live alone, and are most vulnerable to the development and progression of diseases including COVID-19.

In this context, this study describes the main changes in health behaviors, mental and physical health between a pre-pandemic period and weeks 7 to 15 after the beginning the COVID-19 lockdown, among participants in four cohorts of community-dwelling older adults in Spain.

## METHODS

### Study cohorts

#### Seniors-ENRICA-2 (ENRICA)

Longitudinal study of older adults selected between 2015-2017 through sex- and district-stratified random sampling of all community-dwelling individuals aged ≥65 years holding a national healthcare card and living in the Madrid Region (24). In 2019 (baseline wave for the present study), participants were re-interviewed face-to-face at their homes using Computer-Assisted Personal Interviews (CAPI), and underwent a physical exam and blood tests (24). Participants gave informed written consent, and the Clinical Research Ethics Committee of the La Paz University Hospital in Madrid approved the study (Protocol #HULP-PI 1793).

#### Edad con Salud (ES)

Longitudinal population-based household survey representative of the non-institutionalized older adult population aged ≥60 years recruited in 206 municipalities in Spain. The first wave took place between 2011-2012 and was part of the Collaborative Research on Ageing in Europe (COURAGE) study. Participants were re-evaluated twice, between 2014 and 2015, and in 2018 (25). In 2019-2020 (baseline wave for the present study), just before the start of the COVID-19 outbreak, a refreshment sample with participants from 9 municipalities in Barcelona and 7 municipalities in Madrid was added. Interviews were conducted face-to-face by trained lay interviewers at respondents’ homes using CAPI. The research protocol was approved by the Clinical Research Ethics Review Committees of both Parc Sanitari Sant Joan de Déu (Barcelona), and Hospital Universitario La Princesa (Madrid). All participants provided written informed consent for their participation and the treatment of their personal data.

#### Toledo Study for Healthy Ageing (TSHA)

Prospective study of older adults aged ≥65 years recruited in 15 municipalities in the province of Toledo. The cohort was stablished between 2006-2009, and it had follow-up visits in 2011-2013 and 2016-2017 (baseline wave for the present study), when participants were interviewed at their homes using CAPI, underwent a physical exam, and provided blood and urine samples (26). Participants gave informed written consent, and the Clinical Research Ethics Committee of Toledo Hospital Complex approved the study (Protocol # 2203/30/2005).

#### The elderly-Exernet multi-center study (EXERNET)

Multi-center study of non-institutionalized individuals aged ≥65 years recruited in Aragón, Castilla-La Mancha and Madrid. At baseline (2008-2009) and follow-up visits (2011-2012 and 2016-2017 [baseline wave for the present study]) participants responded to a face-to-face questionnaire and underwent a physical exam (27). Participants gave informed written consent, and the Clinical Research Ethics Committee of Aragón approved the study (#18/2008).

### Participants

A total of 4936 participants from the cohorts with at least one pre-pandemic follow-up visit were eligible to participate in the study. From these, 753 individuals could not be contacted, and 717 did not agree to participate. After excluding 120 individuals infected with COVID-19, 97 living with a COVID-19 infected patient, as well as 208 participants with no information on important confounders, a total of 1323 individuals from *ENRICA*, 464 from *ES*, 829 from *TSHA*, and 425 from *EXERNET* comprised the analytical sample (n=3041).

### Study variables

At baseline and at follow-up, we collected data on health behaviors, mental and physical health, and their potential determinants including demographic and social variables, housing conditions, and aging experiences.

#### Baseline information

In all cohorts, baseline information was collected on sociodemographic factors (sex, age, education, civil status); social isolation (cohabitation, frequency of contacts with family and friends); tobacco consumption; diet quality (14-point Mediterranean diet adherence screener questionnaire-MEDAS) (28); physical activity, as per the EPIC-cohort questionnaire (29) in *ENRICA* and the *EEPAQ* questionnaire (30) in *EXERNET*, with results expressed as Metabolic Equivalent Tasks (METs), the Global Physical Activity Questionnaire in *ES* (31), and the Physical Activity Scale for the Elderly (PASE) in the *TSHA*)(32); measured weight and height, with the body mass index (BMI) being calculated as the weight in kg divided by squared height in m; hours of night-time sleep (short sleep defined as ≤6 hours, and long sleep as ≥9 hours); quality of life (the 12-item Short Form (SF) in *ENRICA* (33), the WHO Disability Assessment Schedule 2.0 (WHODAS) in *ES* (34), and the EuroQol-5D (35) in *TSHA* and *EXERNET*; and history of physician-diagnosed chronic conditions (i.e. hypertension, diabetes, osteoarthritis, and depression). Definition of hypertension was based on a self-reported physician diagnosis, current use of anti-hypertensive medication, or a clinical blood pressure reading ≥140/90 mmHg. Type 2 diabetes mellitus was defined as a self-reported physician diagnosis, current use of anti-diabetic medication, or fasting glucose ≥126 mg/d.

Additionally, in some cohorts, information was collected on feelings of loneliness (the 3-item UCLA loneliness scale in *ES*, and a 0 to 5 scale in *ENRICA*)(36); exposure to second-hand smoke (SHS) (*ENRICA*)(37); alcohol intake (all cohorts but *EXERNET*); frequency of snack consumption and use of food to calm anxiety (*ENRICA*); time on sedentary activities (the Nurses’ Health Study questionnaire in *ENRICA*) (38) or overall sedentary time (*ES* and *EXERNET*); hours of daytime sleep (*ENRICA* and *EXERNET*); overall sleep quality (in all cohorts but *TSHA*); other indicators of poor night-time sleep quality (i.e. “difficulty falling asleep”, “awakening during night-time”, “early awakening with difficulty getting back to sleep” and “use of sleeping pills”) and drowsiness (i.e. “being so sleepy at daytime as to need a nap”, “not feeling rested in the morning”, and the Epworth Sleepiness Scale) in *ENRICA* (39); prevalence and characteristics of pain (ENRICA) (40); history of physician-diagnosed coronary heart disease, congestive heart failure, angina or cancer at any site (all cohorts but *EXERNET*), mobility limitations, in *ENRICA* (41); the Mini-Mental State Examination Score (MMSE) (42) (*ENRICA* and *ES*); the Cantril’s Ladder of Life Scale (43) (*ENRICA* and *ES*), and a negative ageing experience scale that ranged from 1 (very positive experience) to 5 (very negative experience) (*ENRICA*).

Information on overall sleep quality was harmonized into a variable with the following categories: “very good”, “good”, “fair”, “poor/very poor.” Moreover, in *ENRICA* a 0 to 7 score with the number of poor sleep quality indicators was constructed as follows: those who answered “sometimes” or “almost always” to the items “difficulty falling asleep”, “awakening during night-time”, “early awakening with difficulty getting back to sleep”, “use of sleeping pills”, “being so sleepy at daytime as to need a nap” or “not feeling rested in the morning”, as well as those with an Epworth Sleepiness Scale score >10, received 1 point; their counterparts received 0 points. Also, in *ENRICA*, a pain scale ranging from 0 (no pain) to 7 (worse pain) was created based on pain frequency, intensity (assessed according to its impact on habitual activities), and number of pain sites. Sporadic (<2 times/week) and frequent (≥2 times/week) pain were assigned scores of 1 and 2, respectively; light, moderate and high intensity pain, scores of 1, 2, and 3, respectively; and 1-2 and ≥3 pain sites scores of 1 and 2, respectively (40).

#### Follow-up information

Phone interviews were performed in the four cohorts between April 27^th^ (week 7 of the pandemic) and June 22^nd^ (week 15), with information being collected on: presence and symptoms of COVID-19 cases within the household; number of outpatient or hospital health care visits during confinement; problems receiving health care or accessing medications during the pandemic; social isolation, degree of social support and responsibilities towards dependents; poor housing conditions (including accessibility problems, noise annoyance, lack of internet access, absence of outdoor views or lack of a terrace/balcony or a private garden/yard to go out during confinement); tobacco consumption; passive tobacco smoke; alcohol drinking; diet quality; grocery modality during confinement (on-line, in-store) and person in charge; eating at a regular time (all cohorts but *ES*); eating more ultra-processed food than usual; eating snacks or food to calm anxiety (*ENRICA*); time spent in physical activities like walking (i.e. to the supermarket, the pharmacy or to walk the dog), exercising at home (i.e. with a stationary bike, a treadmill, fitness devices…), engaging in do-it yourself activities, or doing housework (*ENRICA* and *EXERNET*); the PASE questionnaire (*TSHA* and *EXERNET*); reasons for not doing any physical activity confinement; weight; time on sedentary activities (*ENRICA*) or overall sedentary time (*EXERNET*); hours of night-time sleep; hours of day-time sleep (*ENRICA* and *EXERNET*); poor sleep quality (all cohorts but the TSHA); quality of life (12-SF in ENRICA and WHODAS in *ES*); frequency, intensity and locations of pain (*ENRICA*).

### Statistical analysis

We first evaluated the longitudinal changes in the frequency of smoking, SHS, alcohol intake, diet, physical activity, measured weight, sedentary time, night-time and day-time sleep, sleep quality, overall health and pain between the pre-confinement and confinement periods, by using mixed models with robust standard errors accounting for within-participant correlations induced by repeated measures. These mixed models were fitted with adjustment for sociodemographic, lifestyle and health-related risk factors, including fixed effects for sex, educational level (primary or less, secondary, or university), baseline marital status (married, widowed, single, divorced), and study cohort (model 1); as well as fixed effects for baseline BMI and chronic morbidities, and changes over time in cohabitation status (living alone or not), smoking status (never, former or current), adherence to the Mediterranean diet score (0 to 14 points), physical activity (cohort-specific quartiles), night-time sleep, and overall health (cohort-specific quartiles) (model 2). To reduce the influence of increasing age during follow-up, we also conducted sensitivity analyses by excluding participants whose baseline measures were obtained earlier than 1.5 years before confinement.

In order to identify potential determinants of lifestyle changes or health declines during confinement, we then evaluated the relationship between baseline variables and changes in lifestyles and health outcomes. With this purpose, we used either linear or multinomial regression models that adjusted for baseline socio-demographic factors, housing conditions, smoking status, adherence to the Mediterranean diet, BMI, recreational activity and household activity, night-time sleep, chronic morbidities, time since last follow-up visit, cohort of study, and week of confinement. Although results from these models are given for each predictor and study outcome in tables, we tried to simplify the presentation of results by conducting additional backward stepwise linear and multinomial regression models with the studied baseline predictors (inclusion set at p<0.1). These models were forced to adjust for age and sex, and their results are reported in figures, with red or blue colors identifying subjects with unhealthier and healthier changes, respectively. Categories “a” and “b” from multinomial regression models (defined in the paragraph below) represent these unhealthier (red) and healthier (blue) categories.

Linear (both fully-adjusted and stepwise) regression was used to model the changes in the MEDAS score (all cohorts), weight (all cohorts), recreational and household activities (*ENRICA* and *EXERNET*), the PASE score (*TSHA*), total sedentary time (*ENRICA* and *EXERNET*), specific sedentary activities (*ENRICA*), hours of night-time sleep (all cohorts), number of total poor sleep quality indicators (*ENRICA*), physical component score (PCS) and mental component score (MCS) of the SF-12 (*ENRICA*), the WHODAS-12 (*ES*), and the pain scale (*ENRICA*). Also, multinomial (both fully-adjusted and stepwise) regression was used for the following *a-priori* defined outcomes:

1. Changes in alcohol consumption. Participants in *ENRICA, ES* and *TSHA* cohorts were classified into the following categories: a) drinkers who increased their frequency of consumption (i.e. changed from not daily to daily drinkers); b) drinkers who decreased their frequency of intake (i.e. changed from daily to not daily or to not consumption at all); c) drinkers who did not change their frequency of consumption (reference); and d) not-drinkers who maintained their non-drinking status during confinement.
2. Changes in diet quality. A “worsening” (a) and an “improvement” (b) categories were created including individuals who had either decreased or increased, respectively, their MEDAS score ≥1 point over follow-up; while a “reference” (c) category classified those who experienced no changes or slight increases/decreases (below 1 point) on this score.
3. Changes in weight. Individuals who gained or lost >1 kg were classified into an “increased weight” (a) or “decreased weight” (b) category, respectively, while those who experienced no or small (<1 kg) changes in weight were classified into a “maintained weight” or “reference” (c) category.
4. Changes in physical activity and sedentary time. Participants in *ENRICA, TSHA* and *EXERNET* cohorts were classified as experiencing: a) “Unhealthier changes”, if they decreased physical activity or increased sedentary time more than the observed 75^th^ percentile in each cohort; b) “Healthier changes”, if they decreased activity or increased sedentary time less than the 25^th^ percentile, or if they had increased activity or decreased sedentary time with confinement; and c); “Average” or “reference” changes if they changed between the 25^th^ and 75^th^ percentile.
5. Changes in night-time sleep. The following categories were defined: a) “Worsening”, those who at baseline had normal sleep and developed short or long sleep with confinement; b) “Improvement”, those who at baseline had short or long sleep and developed normal sleep during confinement; and c) “Reference”, those who stayed in the same normal/non-normal sleep category.
6. Changes in sleep quality. Finally, older adults in *ENRICA, ES* and *EXERNET* cohorts were classified into: a) “Worsening”, if they had suffered an increase in the “poor sleep quality” score during confinement; b) “Improvement”, if they decreased in that score, and c) “Reference”, if that had no changes.

Statistical analyses were performed with Stata version 14.1 (StataCorp LP, College Station, TX).

## RESULTS

Supplementary Table 1 shows the prevalence and mean (SD) estimates for the main sociodemographic, lifestyle and health characteristics of participants in the four cohorts at the pre-confinement period (baseline) and the confinement follow-up period. At baseline, the mean age was 74.5 years and the proportion of men 42.3%, although this latter figure varied widely across cohorts (from 21.2% in *EXERNET* to 50.5% in *ENRICA*). The prevalence of university studies also differed across cohorts and ranged from >15% in the *ENRICA* and *ES* to 8% in *TSHAA* and *EXERNET*. Similarly, participants in *ENRICA* and ES showed a higher prevalence of Internet access. Overall, 65% of participants were married, 4 to 10% lived in houses with no outdoor views, and 24 to 37% lacked an outdoor terrace or balcony to go outside during confinement. Regarding social factors, the proportion of participants who lived alone was similar at baseline and during confinement (around 20-25% in all cohorts except for *EXERNET*, were the frequency was 35%). However, the percentage of participants without daily contact with family or friends halved during the pandemic. Moreover, in *ENRICA*, the single cohort where this information was available, no increases in lonely feelings during the study period were observed.

### Changes in tobacco exposure, alcohol intake, diet quality and weight

**Table 1** shows results for changes in the prevalence of smoking, SHS, alcohol intake and diet quality between the pre-confinement and confinement periods. While the prevalence of active smoking remained stable, the likelihood of quitting smoking increased during follow-up. About half of the active smokers at confinement reported no changes in the frequency of tobacco consumption, while 22% reported an increase, and 27% a decrease, in the frequency of smoking since the start of the pandemic. The risk of increased consumption was higher among smokers with bad sleep quality at baseline and was lower among those lacking a terrace or balcony to smoke (**Supplementary Table 2**).

**Table 1:** Longitudinal changes in the prevalence of smoking, second-hand smoke, alcohol intake, diet quality, physical activity, weight, sedentary time, sleep characteristics, overall health and pain between the COVID-19 pre-confinement (t0) and the confinement (t1) periods.

|  | n/total participants for each variable and study cohort |  |  |  | Main analyses |  | Sensitivity analysis |  |
| --- | --- | --- | --- | --- | --- | --- | --- | --- |
|  | ENRICA | ES | TSHA | Exernet | Model 1 | Model 2 | (only participants with time t0-t1<1.5 years) |  |
| Tobacco exposure |  |  |  |  |  |  |  |  |
| Former smokers; RRR (95%CI)<sup>†</sup> | 521/1323 | 139/464 | 193/829 | 71/425 | 1.33 (1.05; 1.66) | 1.26 (1.00; 1.60) | 293/857 | 1.44 (0.94; 2.19) |
| Active smokers; RRR (95%CI)<sup>†</sup> | 118/1323 | 73/464 | 56/829 | 7/425 | 1.10 (0.88; 1.38) | 1.01 (0.80; 1.27) | 106/857 | 1.08 (0.77; 1.50) |
| Exposure to SHS; OR (95%CI)<sup>†</sup> | 70/1197 |  |  |  | 3.64 (1.70; 7.80) | 4.13 (1.95; 8.79) | 16/347 | 1.49 (0.25; 8.88) |
| Alcohol intake; RRR (95%CI)<sup>†</sup> |  |  |  |  |  |  |  |  |
| No daily consumption | 371/1323 | 135/464 | 84/829 |  | 1.03 (0.77; 1.37) | 0.68 (0.54; 0.85) | 263/856 | 0.72 (0.52; 1.00) |
| Daily/almost daily consumption | 389/1323 | 78/464 | 229/829 |  | 1.00 (0.77; 1.31) | 1.17 (0.97; 1.41) | 182/856 | 1.39 (1.03; 1.89) |
| Dietary intake |  |  |  |  |  |  |  |  |
| Snack consumption; OR (95%CI)<sup>†</sup> | 603/1323 |  |  |  | 0.72 (0.60; 0.85) | 0.72 (0.60; 0.85) | 152/392 | 1.09 (0.78; 1.53) |
| Food to calm anxiety; OR (95%CI)<sup>†</sup> | 175/1323 |  |  |  | 0.54 (0.40; 0.73) | 0.56 (0.41; 0.76) | 56/392 | 0.39 (0.21; 0.74) |
| MEDAS index; $\bar{x}$ changes (95%CI)<sup>‡</sup> | 1323 | 464 | 829 | 425 | -0.53 (-0.61; -0.45) | -0.53 (-0.60; -0.45) | 857 | -0.47 (-0.61; -0.34) |
| Adherence to MEDAS items; OR (95%CI)<sup>†</sup> |  |  |  |  |  |  |  |  |
| Olive oil preference item | 1323 | 464 | 829 | 425 | 3.17 (1.67; 6.01) | 4.27 (1.76; 10.36) | 857 | 13.9 (0.20; 99.4) |
| ≥ 4 tablespoon of olive oil/day | 1323 | 464 | 829 | 425 | 0.47 (0.39; 0.57) | 0.51 (0.42; 0.63) | 857 | 0.80 (0.53; 1.21) |
| 2 serving vegetables/day | 1323 | 464 | 829 | 425 | 0.38 (0.28; 0.48) | 0.50 (0.39; 0.65) | 857 | 0.81 (0.32; 2.82) |
| ≥ 2 serving fruit/day | 1323 | 464 | 829 | 425 | 0.70 (0.59; 0.85) | 0.78 (0.63; 0.96) | 857 | 0.60 (0.37; 0.98) |
| < 2 serving red meat/day | 1323 | 464 | 829 | 425 | 1.30 (1.07; 1.82) | 1.61 (1.21; 2.14) | 857 | 0.39 (0.18; 0.87) |
| < 2 serving butter/day | 1323 | 464 | 829 | 425 | 0.98 (0.73; 1.31) | 1.10 (0.81; 1.49) | 857 | 0.21 (0.04; 1.02) |
| <1 soda drink/day | 1323 | 464 | 829 | 425 | 1.20 (0.89; 1.63) | 1.34 (0.97; 1.84) | 857 | 0.59 (0.21; 1.62) |
| ≥7 glasses wine /week | 1323 | 464 | 829 | 425 | 0.65 (0.51; 0.83) | 0.69 (0.53; 0.91) | 857 | 1.29 (0.71; 2.33) |
| ≥ 2 serving legumes/ week | 1323 | 464 | 829 | 425 | 1.49 (1.23; 1.83) | 2.07 (1.63; 2.63) | 857 | 1.57 (1.00; 2.56) |
| ≥ 2 serving fish/week | 1323 | 464 | 829 | 425 | 0.72 (0.60; 0.86) | 0.86 (0.71; 0.99) | 857 | 0.56 (0.35; 0.88) |
| < 2 serving sweets/week | 1323 | 464 | 829 | 425 | 1.24 (1.05; 1.47) | 1.48 (1.23; 1.76) | 857 | 0.85 (0.57; 1.29) |
| ≥ 2 serving nuts/ week | 1323 | 464 | 829 | 425 | 0.97 (0.81; 1.16) | 1.19 (0.96; 1.47) | 857 | 0.91 (0.53; 1.56) |
| poultry >red meet | 1323 | 464 | 829 | 425 | 4.07 (3.26; 5.08) | 5.25 (4.05; 6.80) | 857 | 10.53 (5.47; 20.31) |
| ≥ 2 servings sofrito/week | 1323 | 464 | 829 | 425 | 0.35 (0.29; 0.42) | 0.33 (0.27; 0.40) | 857 | 0.44 (0.30; 0.65) |
| Weight; $\bar{x}$ changes (95%CI)<sup>‡</sup> | 792 | 241 | 313 | 221 | -0.09 (-0.32; 0.14) | -0.57 (-1.29; 0.15) | 845 | -0.57 (-1.29; 0.15) |
| Physical activity; $\bar{x}$ changes (95%CI)<sup>‡</sup> | | | | | | | | |
| Overall Physical activity (METs/h-wk) | 1323 |  |  | 425 | -16.35 (-18.08;-14.6) | -17.48 (-18.61;-16.35) | 857 | -10.70 (-12.08;-9.32) |
| Recreational activity | 1323 |  |  | 425 | -10.95 (-12.02;-9.87) | -11.32 (-12.24;-10.41) | 857 | -10.84 (-12.43;-9.26) |
| Household activity | 1323 |  |  | 425 | -5.39 (-6.66; -4.12) | -6.04 (-7.11; -4.98) | 857 | 0.15 (-1.19; 1.49) |
| PASE score |  |  | 832 |  | -6.42 (-11.14; -1.70) | -16.66 (-19.59;-13.73) | - | - |

|  | n/total participants for each variable<br>and study cohort |  |  |  | Main analyses |  | Sensitivity analysis |  |
| --- | --- | --- | --- | --- | --- | --- | --- | --- |
|  | ENRICA | ES | TSHA | Exernet | Model 1 | Model 2 | (only participants with time t0-t1<1.5 years) |  |
| **Sedentary time; $\bar{x}$ changes (95%CI) ‡** |  |  |  |  |  |  |  |  |
| **Overall** (min/day) | 1323 |  |  | 425 | **91.89 (81.61; 102.1)** | **91.68 (81.21; 102.11)** | 393 | **70.53 (49.99; 91.17)** |
| **TV time** (min/day) | 1323 |  |  |  | **27.18 (19.56; 34.80)** | **27.93 (20.23; 35.63)** | 393 | **24.07 (10.12; 38.02)** |
| **Other screen time** (min/ day) | 1319 |  |  |  | **50.20 (45.57; 54.83)** | **50.60 (45.83; 55.44)** | 393 | **53.99 (45.32; 62.64)** |
| **Reading** (min/d) | 1323 |  |  |  | **16.30 (12.34; 20.25)** | **15.53 (11.37; 19.58)** | 393 | **14.97 (7.68; 22.28)** |
| **Listening to music** (min/d) | 1323 |  |  |  | **7.52 (4.61; 10.43)** | **7.25 (4.26; 10.24)** | 393 | **6.94 (1.77; 12.12)** |
| **Sleep characteristics; $\bar{x}$ changes (95%CI) ‡** |  |  |  |  |  |  |  |  |
| **Hours /night-time sleep** | 1323 | 464 | 829 | 425 | -0.02 (-0.08; 0.04) | -0.00 (-0.06; 0.05) | 857 | **-0.16 (-0.27; -0.06)** |
| **Minutes/day-time sleep** | 1317 |  |  | 421 | **-12.50 (-14.18;-10.8)** | **-12.08 (-13.73;-10.43)** | 393 | **-8.70 (-11.84; -5.56)** |
| **Poor sleep quality score** | 1289 | 430 |  | 395 | **0.13 (0.10; 0.16)** | **0.12 (0.09; 0.15)** | 857 | **0.14 (0.09; 0.19)** |
| **N° poor sleep indicators** | 1285 |  |  |  | 0.06 (-0.02; 0.14) | **0.08 (0.00; 0.15)** | 390 | 0.04 (-0.09; 0.18) |
| **Overall health§** |  |  |  |  |  |  |  |  |
| **SF-12, PCS; $\bar{x}$ changes (95%CI) ‡** | 1311 |  |  |  | **5.03 (4.45; 5.60)** | **4.79 (4.72; 5.38)** | 389 | **4.57 (3.45; 5.71)** |
| **SF-12, MCS; $\bar{x}$ changes (95%CI) ‡** | 1311 |  |  |  | **-1.20 (-1.80; -0.58)** | **-1.19 (-1.81; -0.58)** | 389 | **-1.41 (-2.48; -0.37)** |
| **WHODAS-12; $\bar{x}$ changes (95%CI) ‡** |  | 464 |  |  | **2.57 (1.29; 3.84)** | -0.18 (-1.85; 1.48) | 464 | -0.18 (-1.85; 1.48) |
| **Pain scale; $\bar{x}$ changes (95%CI) ‡** | 1296 |  |  |  | **0.17 (0.06; 0.30)** | **0.18 (0.05; 0.30)** | 323 | **0.27 (0.05; 0.49)** |

Note: Figures highlighted in bold show a statistically significant association.
ENRICA: Seniors-ENRICA-2. ES: Edad con Salud. TSH: Toledo Study for Healthy Ageing.
$\bar{x}$ : Mean change. OR: Odds Ratio; 95%CI: 95% Confidence interval; MEDAS: Mediterranean Diet Assessment Score; Met: Metabolic Equivalent to Task; MCS: Mental Component Score of the SF-12; PA: Physical Activity; PCS: Physical Component Score of the SF-12; rPA:; SHS: Second-hand smoke.
† Odds ratios and Relative Risk Ratios (RRR) (95% confidence intervals) for tobacco smoke exposure, alcohol intake, snack consumption, intake of food consumption to calm anxiety, short- or long-sleep duration and poor sleep quality between the pre-confinement and confinement periods were obtained from multilevel mixed effects logistic (OR) or multilevel structural equation models (RRR), with robust standard errors accounting for within-participant correlations induced by repeated measures.
‡ Average differences (95% confidence intervals) in MEDAS, weight, physical activity, sedentary time, hours of night- or day-time sleep, and number of poor sleep quality indicators between the pre-confinement and confinement periods were obtained from repeated measures linear regression models with robust standard errors accounting for within-participant correlations induced by repeated measures.
§ Higher scores in the mental component and physical component of the SF-12 and lower scores in the WHODAS-12 are indicative of better health.
Model 1 adjusted for age, sex (men or women), educational level (primary or less, secondary, or university), civil status (married, widowed, single or divorced) and study cohort. Model 2 further adjusted for baseline body mass index (normoweight, overweight or obese) and chronic comorbidities (hypertension, diabetes, osteo-muscular disease and depression); as well as for changes over time in cohabitation (living alone, not living alone), smoking (never, former or current), adherence to the Mediterranean diet (MEDAS score), physical activity (cohort-specific quartiles), hours of night-time sleep (normal, short sleep or long sleep), and overall health (in quartiles).

**Table 2:** Prospective association between participant characteristics and changes in the mental component (MCS) and physical component (PCS) scores of the SF-12 (n=1243 from *Seniors-ENRICA-2*), the WHODAS-12 (n=460 from *Edad con Salud*), and the pain score (n=983 from *Seniors-ENRICA-2*) during the COVID-19 confinement. Increases in MCS or PCS components reflect improvements in health, while increases in the WHODAS-12 or pain scores reflect worsening in health or pain, respectively.

|  | Changes in health |  |  |  |
| --- | --- | --- | --- | --- |
|  | Changes in PCS | Changes in MCS | Changes in WHODAS-12 | Changes in pain scale<sup>†</sup> |
| | $\bar{x}$ (95%CI)<br>(n=1243) | $\bar{x}$ (95%CI)<br>(n=1243) | $\bar{x}$ (95%CI)<br>(n=460) | $\bar{x}$ (95%CI)<br>(n=983) |
| **Socio-demographic characteristics** |  |  |  |  |
| Age, yr | **-0.19 (-0.29; -0.08)** | **0.12 (0.00; 0.23)** | 0.14 (-0.05; 0.33) | 0.01 (-0.01; 0.04) |
| Female | -0.16 (-1.21; 0.89) | **-2.46 (-3.65; -1.35)** | -0.93 (-3.73; 1.88) | **0.28 (0.03; 0.51)** |
| **Education** |  |  |  |  |
| Primary or less | Ref. | Ref. | Ref. | Ref. |
| Secondary | -0.17 (-1.28; 0.93) | 0.18 (-1.02; 1.38) | -0.63 (-3.57; 2.32) | -0.05 (-0.31; 0.21) |
| University | 0.90 (-0.20; 1.99) | 0.11 (-1.08; 1.31) | **-4.12 (-7.70; -0.53)** | -0.13 (-0.30; 0.12) |
| **Civil status** |  |  |  |  |
| Married | Ref. | Ref. | Ref. | Ref. |
| Single | -0.74 (-2.74; 1.25) | **2.16 (0.00; 4.33)** | 2.68 (-3.82; 9.28) | 0.29 (-0.19; 0.76) |
| Divorced | -0.02 (-2.05; 2.01) | 0.94 (-1.26; 3.15) | 1.80 (-2.78; 6.39) | 0.20 (-0.31; 0.70) |
| Widowed | -0.75 (-2.20; 0.69) | 1.24 (-0.33; 2.81) | -1.79 (-6.04; 2.45) | 0.14 (-0.21; 0.49) |
| Living alone | 0.74 (-0.72; 2.20) | **-1.40 (-3.00; 0.09)** | -0.10 (-4.06; 3.87) | -0.12 (-0.48; 0.23) |
| No daily contact with family/friends other than cohabitants | 0.23 (-1.04; 1.50) | -0.24 (-1.62; 1.14) | 0.46 (-2.24; 3.17) | 0.06 (-0.05; 0.18) |
| Feeling lonely | 0.22 (-0.26; 0.70) | **-1.02 (-1.54; -0.50)** | 0.37 (-0.78; 1.52) | 0.06 (-0.05; 0.18) |
| **Housing conditions** |  |  |  |  |
| Lack of outdoor views | 0.40 (-1.12; 1.92) | -0.94 (-2.60; 0.71) | 3.29 (-0.93; 7.51) | 0.14 (-0.22; 0.50) |
| Lack of terrace/balcony | 0.27 (-0.69; 1.21) | 0.23 (-0.80; 1.27) | -1.95 (-4.56; 0.66) | -0.05 (-0.28; 0.18) |
| Lack of garden/yard | **-1.38 (-2.60; -0.16)** | -0.44 (1.78; 0.89) | -1.09 (3.77; 1.60) | 0.02 (-0.28; 0.18) |
| No internet access | 0.45 (-0.66; 1.56) | -0.50 (-1.70; 0.70) | 1.61 (-1.62; 4.84) | -0.02 (-0.28; 0.24) |
| Too much noise | 1.62 (-1.18; 4.42) | **-5.05 (-8.10; -2.00)** | -0.00 (-5.80; 5.79) | **0.74 (0.10; 1.38)** |
| **Lifestyle-behaviors** |  |  |  |  |
| **Smoking** |  |  |  |  |
| Never smokers | Ref. | Ref. | Ref. | Ref. |
| Former smokers | 0.12 (-0.83; 1.08) | 0.22 (-0.81; 1.26) | -1.93 (-4.88; 1.03) | 0.05 (-0.17; 0.27) |
| Smoker | 0.10 (-1.42; 1.61) | -0.62 (-2.27; 1.03) | 0.95 (-2.73; 4.62) | -0.13 (-0.48; 0.22) |
| **Alcohol intake** |  |  |  |  |
| Not drinker | Ref. | Ref. | Ref. | Ref. |
| Drinker, not daily | -0.05 (-1.08; 0.99) | -0.23 (-1.36; 0.90) | -0.02 (-2.92; 2.88) | 0.03 (-0.21; 0.28) |
| Drinker, daily/almost daily | -0.15 (-2.22; 0.92) | -0.33 (-1.49; 0.84) | -0.94 (-4.60; 2.72) | 0.08 (-0.17; 0.33) |
| **MEDAS** | -0.02 (-0.27; 0.23) | 0.11 (-0.16; 0.38) | -0.28 (-0.98; 0.42) | 0.06 (-0.00; 0.12) |
|  | Changes in PCS | Changes in MCS | Changes in WHODAS-12 | Changes in pain scale<sup>†</sup> |
| | $\bar{x}$ (95%CI)<br>(n=1243) | $\bar{x}$ (95%CI)<br>(n=1243) | $\bar{x}$ (95%CI)<br>(n=460) | $\bar{x}$ (95%CI)<br>(n=983) |
| **PA (cohort-specific quartiles)** |  |  |  |  |
| 1st quartile (less) | Ref. | Ref. | Ref. | Ref. |
| 2nd quartile | 0.58 (-0.59; 1.75) | -0.74 (-2.01; 0.53) | -2.64 (-6.07; 0.79) | 0.06 (-0.22; 0.33) |
| 3rd quartile | 1.61 (0.40; 2.83) | -1.05 (-2.34; 0.28) | **-5.48 (-8.92; -2.02)** | -0.12 (-0.40; 0.16) |
| 4th quartile | 1.03 (-0.25; 2.31) | **-1.35 (-2.75; 0.05) \*** | **-4.20 (-7.90; -0.49) \*** | -0.02 (-0.32; 0.28) |
| **BMI** |  |  |  |  |
| Normoweight | Ref. | Ref. | Ref. | Ref. |
| Overweight | -0.11 (-1.10; 0.89) | 0.05 (-1.03; 1.13) | -1.23 (-4.20; 1.75) | -0.14 (-0.37; 0.09) |
| Obese | -0.14 (-1.38; 1.10) | -0.29 (-1.64; 1.06) | -1.18 (-4.47; 2.10) | -0.05 (-0.34; 0.24) |
| **ST (cohort-specific quartiles)** |  |  |  |  |
| 1st quartile (less time) | Ref. | Ref. | Ref. | Ref. |
| 2nd quartile | -0.31 (-1.48; 0.86) | -0.55 (-1.83; 0.71) | 0.32 (-3.05; 3.68) | 0.24 (-0.04; 0.51) |
| 3rd quartile | -0.26 (-0.89; 1.41) | -1.06 (-2.31; 0.19) | -0.39 (-3.62; 2.85) | -0.06 (-0.33; 0.21) |
| 4th quartile | -0.53 (-1.71; 0.64) | -0.16 (-1.43; 1.12) | 3.39 (-0.38; 7.15) | -0.03 (-0.31; 0.25) |
| **Sleep characteristics** |  |  |  |  |
| **Hours of night-time sleep** |  |  |  |  |
| Normal sleep | Ref. | Ref. | Ref. | Ref. |
| Short sleep ($\leq 6$ h) | -0.62 (-1.76; 0.51) | -0.04 (-1.27; 1.19) | 1.03 (-2.65; 4.70) | **0.46 (0.19; 0.73)** |
| Long sleep ($\geq 9$ h) | 0.67 (-1.34; 2.68) | -1.32 (-3.51; 0.87) | -1.76 (-5.62; 2.11) | -0.29 (-0.76; 0.17) |
| **Hours of day-time sleep<sup>‡</sup>** |  |  |  |  |
| None | Ref. | Ref. | - | Ref. |
| Short nap ($\leq 30$ min) | 0.04 (-0.93; 1.01) | -1.15 (-2.20; -0.10) | - | -0.11 (-0.34; 0.11) |
| Long nap (30-60 min) | -0.81 (-2.08; 0.46) | 0.50 (-0.87; 1.87) | - | -0.04 (-0.34; 0.25) |
| Very long nap ($\geq 60$min) | 1.26 (-1.74; 2.27) | **-2.49 (-4.66; -0.33)** | | -0.19 (-0.66; 0.27) |
| **Overall sleep quality** |  |  |  |  |
| Very good | -0.47 (-1.81; 0.87) | 2.09 (0.64; 3.53) | -0.58 (-3.20; 2.05) | 0.04 (-0.27; 0.35) |
| Good | Ref. | Ref. | Ref. | Ref. |
| Fair | -0.65 (-1.73; 0.44) | **-1.92 (-3.01; -0.75)** | 3.15 (-1.38; 7.68) | 0.18 (-0.08; 0.43) |
| Poor/very poor | **-4.04 (-7.07; -1.02) \*** | 0.49 (-2.78; 3.77) | -3.79 (-16.7; 9.10) | -0.04 (-0.83; 0.75) |
| **Health-related variables** |  |  |  |  |
| **Overall health** |  |  |  |  |
| 1<sup>st</sup> quartile | - | - | - | Ref. |
| 2<sup>nd</sup> quartile | - | - | - | -0.46 (-0.80; -0.13) |
| 3<sup>rd</sup> quartile | - | - | - | -0.56 (-0.92; -0.22) |
| 4<sup>th</sup> quartile (best) | - | - | - | -0.64 (-1.00; -0.27) † |
|  | Changes in PCS | Changes in MCS | Changes in WHODAS-12 | Changes in pain scale<sup>†</sup> |
| | $\bar{x}$ (95%CI)<br>(n=1243) | $\bar{x}$ (95%CI)<br>(n=1243) | $\bar{x}$ (95%CI)<br>(n=460) | $\bar{x}$ (95%CI)<br>(n=983) |
| **Chronic morbidities** |  |  |  |  |
| Diabetes | 0.56 (-0.54; 1.67) | -0.22 (-1.42; 0.30) | 0.32 (-3.02; 3.67) | -0.05 (-0.32; 0.22) |
| Hypertension | **-0.22 (-1.08; 0.64)** | -0.55 (-1.49; 0.39) | -0.30 (-2.83; 2.23) | 0.05 (-0.15; 0.25) |
| CVD<sup>‡</sup> | **-2.53 (-4.72; -0.34)** | 0.98 (-1.41; 3.36) | -0.59 (-4.52; 3.33) | 0.24 (-0.22; 0.49) |
| Cancer<sup>‡</sup> | **-1.98 (-3.76; -0.20)** | -0.81 (-2.75; 1.13) | 0.84 (-3.21; 4.89) | 0.19 (-0.27; 0.66) |
| Osteomuscular disease | -0.69 (-1.58; 0.21) | 0.33 (-0.65; 1.30) | 2.49 (-0.13; 5.10) | **0.57 (0.36; 0.79)** |
| Depression | **-1.87 (-3.39; -0.34)** | -1.36 (-3.91; 0.30) | 0.88 (-4.50; 13.05) | **0.30 (0.00; 0.65)** |
| **Mobility limitations** | **-2.37 (-3.49; 1.25)** | **-1.67 (-2.90; -0.44)** |  | **0.25 (0.01; 0.50)** |
| **Negative ageing experience scale** | -0.16 (-0.58; 0.26) | **-0.85 (-1.31; -0.40)** |  | 0.02 (-0.08; 0.12) |
| **Cantril ladder** | **0.28 (0.03; 0.52)** | 0.20 (-0.06; 0.47) |  | **-0.11 (-0.16; -0.05)** |
| **Low MMSE score (<23)** | 1.59 (-1.16; 4.34) | **-3.24 (-6.23; -0.26)** |  | 0.18 (-0.52; 0.88) |
| **Years since baseline examination** | -0.50 (-1.46; 0.47) | 0.48 (-0.57; 1.54) | 0.50 (-1.46; 0.47) | **-0.23 (-0.46; -0.00)** |
| **Week of the state of alarm** | **-0.64 (-1.07; -0.20)** | **0.99 (0.52; 1.46)** | 0.39 (-0.67; 1.45) | **0.10 (0.00; 0.20)** |

$\bar{x}$ : Mean change. 95%CI: 95% Confidence interval; MEDAS: Mediterranean Diet Assessment Score; PA: Physical activity; BMI: Body Mass Index; ST: Sedentary time; PCS: Physical Component Score of the SF-12; MCS: Mental Component Score of the SF-12; MMSE: Mini Mental State Examination
Note: Figures highlighted in bold show a statistically significant association.
<sup>†</sup> The pain scale ranged 0 (no pain) to 7 (worst pain) and was created based on pain frequency, pain intensity (assessed according to its impact on habitual activities), and number of pain sites. Sporadic (<2 times/week) and frequent ( $\geq 2$ times/week) pain were assigned a score of 1 and 2, respectively; light, moderate and high intensity, a score of 1, 2, and 3, respectively; and 1-2 and $\geq 3$ sites a score of 1 and 2, respectively.
<sup>‡</sup> Information only available in the *Seniors ENRICA-2* cohort
\* p-value for trend <0.05
All models were adjusted for baseline age, sex (men or women), educational level (primary or less, secondary, or university), civil status (married, widowed, single, divorced), housing conditions (lack of outdoor views, terrace/balcony, private garden/yard, lack of internet and noise), smoking status (never, former, or current), adherence to the Mediterranean diet (MEDAS score), body mass index (normoweight, overweight, or obese), physical activity (cohort-specific quartiles), hours of night-time sleep (normal, short sleep, long sleep), chronic comorbidities (hypertension, diabetes, osteo-muscular disease and depression), time since last follow-up visit, cohort of study and week of the State of Alarm. Models for MCS and PCS components of the SF- 12 adjusted for baseline values of the PCS and the MCS, respectively.

In *ENRICA*, the prevalence of SHS increased during follow-up, although this association was lost when excluding participants who had been followed-up for more than 1.5 years. The prevalence of current alcohol drinking was very similar in both periods, whereas a decreased risk of not daily alcohol consumption, and a non-significant increased risk of daily intake, were observed. Despite this change in the frequency of consumption, data from *ENRICA* showed that the prevalence of binge drinking (defined as having >5 units of alcohol in a session in men, or >4 in women) was lower during confinement (1.8%) than before (2.7%) (data not shown in tables). Among drinkers, only a few baseline factors were associated with the odds of changing the frequency of alcohol consumption; notably participants who lacked a terrace or balcony and former smokers showed a lower odds of increasing their frequency of alcohol intake during follow-up (**Figure 1** and **Supplementary table 2**).

**Figure 1:**
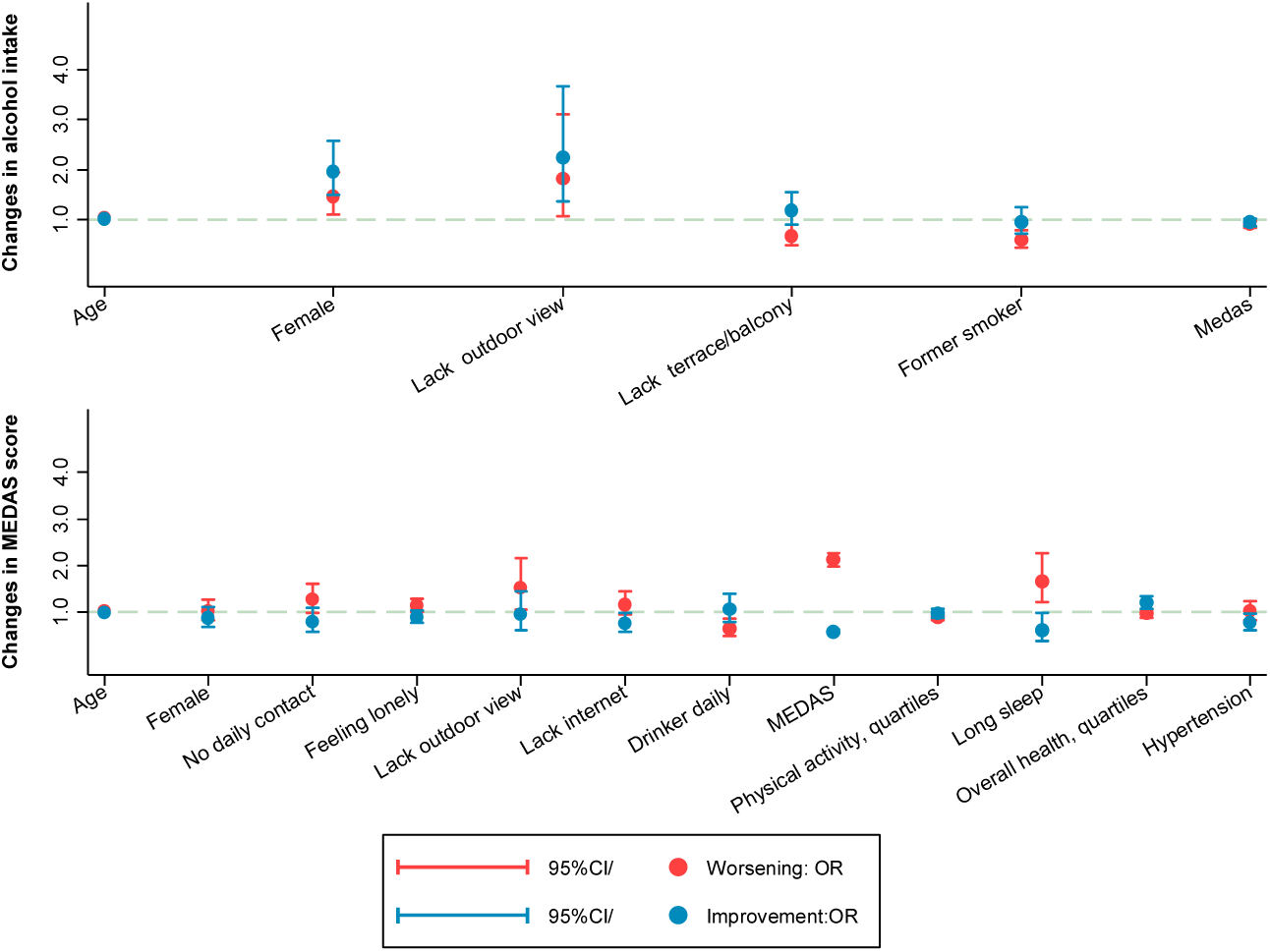
Prospective association between baseline participant characteristics and changes in alcohol consumption or in adherence to the Mediterranean diet adherence screener (MEDAS) score during confinement. Worsening was defined by increasing alcohol frequency from not daily to daily, or by decreasing the MEDAS score in one or more points during follow-up. Improvement was defined by decreasing alcohol frequency from daily to not daily, or by increasing the MEDAS scores in one or more points during follow-up. Variables were selected with a backward stepwise regression from the list of candidate variables shown in **<u>Supplementary</u> <u>Tables</u> <u>2-3</u>**.

Most study participants reported eating at a regular time during confinement, and a very low proportion declared eating more ultra-processed food than before (**Supplementary Table 1**). During confinement, 51.5% of study participants shopped face-to-face, 41.7% asked someone to shop for them, and 6.8% shopped online (data not shown in tables). In *ENRICA*, the odds of snacks consumption and that of use of food to calm anxiety was lower during the State of Alarm than in the pre-confinement period ([odds ratio (OR): 0.72; 95%CI: 0.60; 0.85] and [OR: 0.56; 95%CI: 0.41; 0.76]), respectively (**Table 1**), although the association with snacks consumption was lost when excluding participants whose follow-up was longer than 1.5 years. Data from all cohorts combined showed a mean reduction of 0.5 points (95%CI: -0.60; -0.45) in the MEDAS score during follow-up (**Table 1**). The items that were most negatively affected by the State of Alarm were fruits and vegetables, fish, wine, olive oil and *sofrito* (a sauced made of olive oil and garlic); while the preference for olive oil and red meat, consumption of legumes and consuming less sweets and cakes, improved. Predictors of increased odds of worsening diet quality with confinement included having better MEDAS scores at baseline, being older, lacking outdoor views, and sleeping > 9 hours/night (**Figure 1** and **Supplementary table 3**). A lower odd of worsening diet quality was observed for daily moderate drinkers and for individuals who performed more physical activity, while those with better general health and non-hypertensives were more likely to experience improvements in the MEDAS. No differences in changes in the MEDAS score were observed by grocery modality (data not shown).

Although 29% and 19% of participants reported “feeling they were getting fatter or thinner” with confinement, respectively, no changes in weight between study periods were observed in those with objective information (mean change: -0.57 kgs; 95%CI: - 1.29; 0.15). Still, self-reported and objective information showed some correlation: on average, those who self-reported “feeling they were getting fatter” had gained 1.3 kg since the pre-confinement measure was taken, while those who self-reported “feeling they were losing weight” had lost, on average, -2.3 kg. Women, those with university studies, short night-time sleepers and those who took longer naps at baseline, experienced greater weight loss during follow-up than their counterparts (**Supplementary table 4)**.

### Changes in physical activity and sedentary time

Physical activity and sedentary time were the lifestyle behaviors that changed the most with confinement (**Table 1 and Supplementary Table 1**). Compared to the pre-confinement period, participants in *ENRICA* and *EXERNET* showed a mean reduction in recreational and household physical activity of 17.48 METs/h-week (95%CI: -18.61; - 16.35) and 11.32 METs/h-week (95%CI: -12.24; -10.41), respectively; participants in *TSHA* showed a mean reduction in the PASE score of 16.66 points (95%CI: -19.59; - 13.73). Alleged reasons for not doing any physical activity during confinement among those who used to engage in some type of recreational activity before confinement included not being able to go outside (52.4%), not knowing how to perform physical activity at home (11.4%), not having the resources to do physical activity at home (13.3%), or not having enough time (3.4%).

Regarding sedentary time, the *ENRICA* and *EXERNET* cohorts showed an average increase of 91.68 minutes/day (95%CI: 81.21; 102.11), which, according to the *ENRICA*, was more marked for active (i.e. time on the computer or tablet, time reading) than passive (i.e. television viewing) sedentary activities.

Interestingly, changes in physical activity and sedentary time seemed to reverse with time. Compared to those interviewed in the first weeks of total confinement, those interviewed when confinement measures started to be lifted showed a progressive increase in activity and a decrease in sedentary time: average changes per 1 further week of the State of Alarm were 1.45 METs/h-week (95%CI: 0.24; 2.66) for recreational activity, -0.30 minutes/day (−0.44; -0.15) for sedentary time, and 8.83 points (95%CI: 4.31; 13.4) for the PASE score (**Figure 2** and **Supplementary Tables 5 and 6)**.

**Figure 2.**
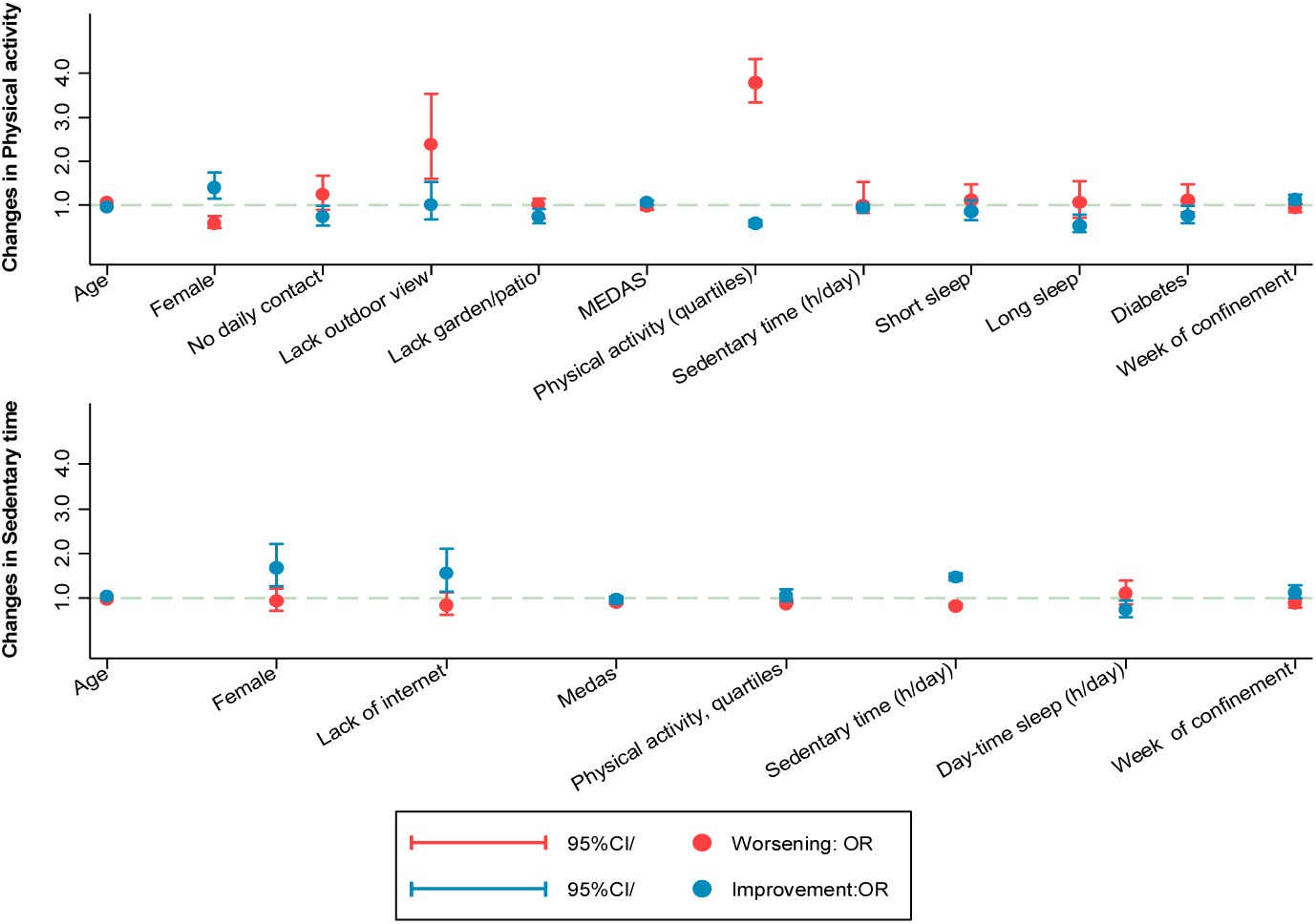
Prospective association between baseline participant characteristics and changes in physical activity and sedentary time during confinement. Worsening was defined by decreasing physical activity or increasing sedentary time more than the observed 75^th^ percentile change. Improvement was defined by decreasing or increasing sedentary time less than observed 25^th^ percentile change Variables were selected with a backward stepwise regression from the list of candidate variables shown in **<u>Supplementary</u> <u>Tables</u> <u>5-6</u>**.

Compared to men, women showed higher odds of presenting healthier changes in activity (OR: 1.34; 95%CI: 1.03; 1.74) and sedentary time (OR: 1.54; 95%CI: 1.08; 2.19), as they seemed to compensate the inability to perform some recreational activities during confinement with increases in household activity; as well as higher reductions in TV viewing time (−0.31 minutes/day; 95%CI: -0.60; -0.02) and higher increases in reading time (0.19 minutes/day; 95%CI: 0.03; 0.45). In general, poor housing conditions like not having outdoor views, or lacking a garden/yard were associated with unhealthier changes in physical activity, as well as with higher increases in TV time with confinement; while not having internet access was associated with lower increases in sedentary time, particularly in screen time. Participants with higher baseline MEDAS scores were less likely to experience unhealthy changes in physical activity or sedentary time, while those who performed more activity were less likely to increase sitting time, and those who spent more time seated were less likely to reduce physical activity. Poor night-time sleep duration and daytime sleepiness at baseline were also associated with unhealthier changes in activity and sedentary time.

### Changes in sleep duration and sleep quality

In multivariate models adjusted for changes in other potential confounders, data from all cohorts combined showed no changes in night-time sleep duration with confinement (−0.00 hours/night; 95%CI: 0.06; 0.05). However, slight decreases were observed in day-time sleep duration (−12.08 minutes/day; 95%CI -13.73, -10.42), as well as in the “poor sleep quality” scale (0.12 points; 95%CI: 0.09; 0.15) (**Table 1**). Baseline predictors of unhealthier changes in sleep quality included being a married woman, living alone, feeling lonelier, doing more physical activity, a lower adherence to the Mediterranean diet, having worse overall health and greater pain (**Supplementary Table 7** and **Figure 3**).

**Figure 3.**
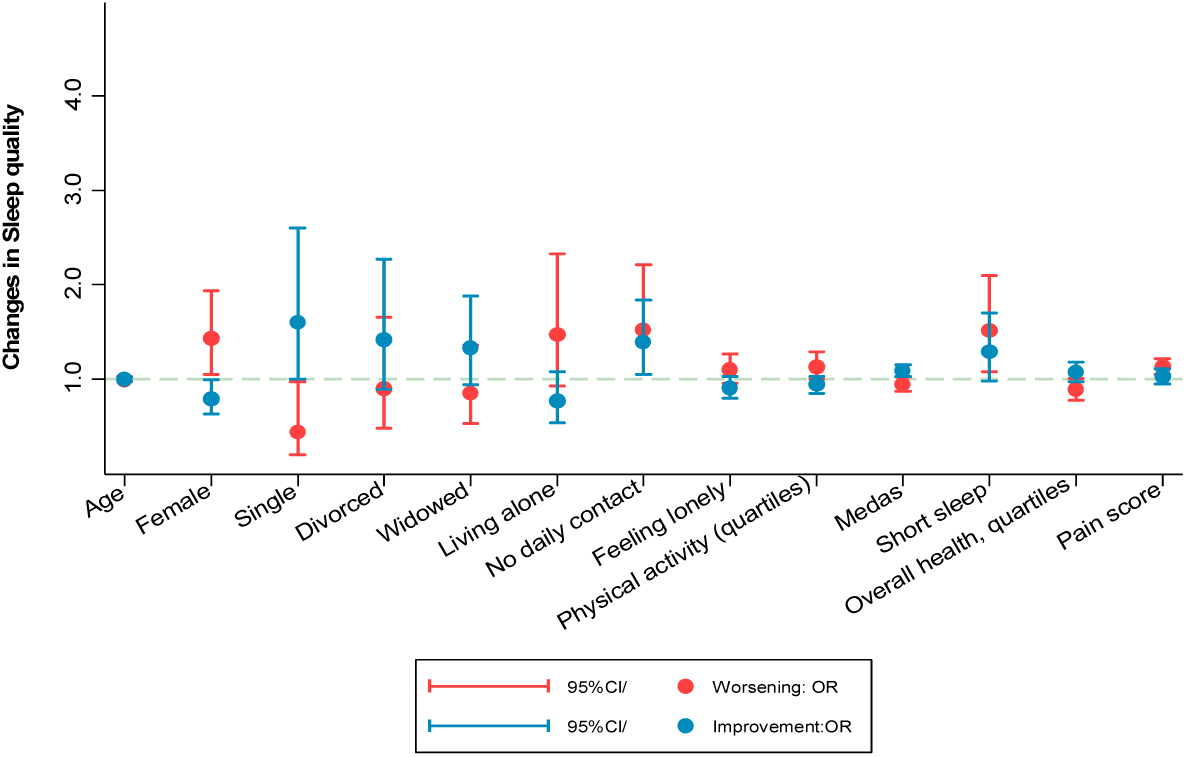
Prospective association between baseline participant characteristics and changes in poor sleep quality score during confinement. Worsening was defined as an increase in the poor sleep quality score, and improvement as a decrease in the same score. Variables were selected with a backward stepwise regression from the list of candidate variables shown in **<u>Supplementary</u> <u>Table</u> <u>7</u>**.

### Changes in physical and mental health and pain

During confinement, 23% and 4.9% of study participants attended a primary health center or an emergency room for reasons not related to a COVID-19 infection. Most of them (87.4% and 91.2%, respectively) declared they were satisfied with the care received. Most participants took their usual medication, although a few (n=4) declared not being able to get medications because of the imposed mobility restrictions (data not shown in tables).

In multivariate analyses, participants in *ENRICA* showed a mean increase of 4.79 points (95%CI: 4.72; 5.38) in the PCS of the SF-12, a reduction of 1.19 points (95%CI: -1.81; - 0.58) in the MCS of the SF-12, and an increase of 0.18 points (95%CI: 0.05-0.30) in the pain scale (**Table 1**); while the participants in the *ES* study showed a non-significant 0.18-point decrease (95%CI: -1.85; 1.48) in the WHODAS score. **Table 2** shows the main predictors of health decline during confinement. Interestingly, the positive changes in the PCS reverted when participants returned to their normal activities (mean changes in the PCS per week of the State of Alarm: -0.64; 95%CI: -1.07; -0.20), while the effects on mental health (mean changes per week of State of Alarm: 0.99; 95%CI: 0.52; 1.46) and pain (0.10; 95%CI: 0.00; 0.20) showed a stronger magnitude in those interviewed later in time. As shown in **Table 2**, older adults experienced unhealthier changes in the PCS (mean changes per year increase: -0.19; 95%CI: -0.29; -0.08) and the WHODAS-12 (mean changes per year increase: 0.14; 95%CI:-0.05; 0.33), but healthier changes in the MCS (mean changes per year increase: 0.12; 95%CI: 0.00; 0.23). Women in *ENRICA* showed greater deteriorations in mental health (mean changes in MCS: -2.46; 95%CI: - 3.65; -1.35) and pain (mean changes in pain score: 0.28; 95%CI:0.03; 0.51), while participants with higher education in the *ES* study showed healthier changes in the WHODAS-12 (mean change -4.12; 95%CI: -7.70; -0.53), than their counterparts.

Other predictors of unhealthier changes in the PCS were lacking a garden/yard, poorer sleep quality, hypertension, cardiovascular disease, cancer, depression, mobility limitations, and lower general life-satisfaction. Predictors of unhealthier changes in MCS were living alone, feeling lonelier, having too much noise at home, doing less physical activity, taking very long naps, having mobility limitations, a negative ageing experience, and low MMSE scores. Finally, unhealthier changes in pain were observed in participants who lived in homes with more noise, as well as in those who suffered from baseline short sleep, worse overall health, osteo-muscular disease, depression, mobility limitations or lower general life-satisfaction.

### Sensitivity analyses

When analyses were restricted to the 857 participants whose baseline measurements had been taken as far apart as 1.5 years before the State of Alarm, consistent findings were observed for most variables except for SHS, snacks consumption, and household physical activity, which no longer showed significant changes. The relationship with changes in some MEDAS items (i.e. vegetables, red meat, wine, sweets and cakes) also disappeared, and a slight decrease in night-time sleep was now observed (mean change -0.16 hours/day; 95%CI: -0.27; -0.06).

## DISCUSSION

This study, conducted with older adults from four Spanish population-based cohorts, showed that, on average, strict confinement was not associated with a deterioration in lifestyle risk factors, except for physical activity and sedentary time, whose effects seemed to reverse with the end of the State of Alarm. Despite this, results from the *ENRICA* suggested moderate declines in mental health that did not seem to reverse after restrictions were lifted. On the other hand, the apparent improvements in physical health in participants from this last cohort did reverse with the end of the State of Alarm and may have been related to a better self-perceived performance at a less physically demanding time.

This study identified subgroups of individuals at increased risk of developing unhealthier lifestyles with confinement, including males (for physical activity and sedentariness), those without daily contact with family and friends (for diet and physical activity), married participants (for sleep quality), those with feelings of loneliness (for diet and sleep quality), those with poor housing conditions (for diet, physical activity, TV viewing time), those with unhealthy sleep duration, as well as those with worse overall health or chronic morbidities (i.e., subjects with obesity or diabetics suffered greater reductions in physical activity and greater increases in screen time). On the other hand, having a greater adherence to the Mediterranean diet and doing more physical activity protected older adults from developing unhealthier lifestyles with confinement.

### What do other studies say about changes in smoking, alcohol intake, diet quality and weight during the COVID-19 pandemic?

Different on-line surveys conducted during the first months of the pandemic in adults from China (44), France (45), the Netherlands (46), or the United Kingdom (47), have shown that current smokers were similarly likely (around 20-25%) to increase or decrease their tobacco consumption with the pandemic. In the Dutch survey, authors found that this apparently contradictory relationship was driven both by a negative effect of stress, boredom and restriction on smoking, and by a positive effect of motivation to quit smoking from fearing to contract COVID-19 and become severely ill. Consistently, in a Chinese survey, participants with higher epidemiologic concern were more often willing to improve their diets (48). In the UK study, on the other hand, symptoms of mental distress (i.e. anxiety, low mood, sleeping problems) during quarantine were more frequently associated with increasing than decreasing tobacco smoke. Although in secondary analysis we did not find a relationship between symptoms of psychological distress and depression (measured at follow-up with the General Health Questionnaire-12 (49) and the Geriatric Depression Scale (50)) and changes in smoking frequency, we did observe that smoking older adults who spent more time listening or reading news related to the COVID-19 outbreak were more likely to increase their tobacco consumption (data not shown in tables). Our study provides evidence that smokers with poor sleep quality may be at higher risk of increasing smoking frequency in stressful situations than those who have good sleep quality. Also, it shows that while “lacking a terrace or balcony” was positively associated with unhealthier changes in physical activity and sedentary time with confinement, it reduced the risk of smokers and drinkers to increase their frequency of tobacco and alcohol intake, respectively.

In our study, the frequency of alcohol drinkers who increased and that of those who decreased alcohol intake with lockdown was very similar, around 15%. Compared to other surveys, the frequency of increased alcohol consumption was much lower in our population than in adults in Germany (34.7%) (51), Australia (20.0% (52) to 30.8% (53)) or the USA (29%) (54), and more similar to that reported in other Spanish (14,22), French (45,55), and Italian surveys (56). In fact, in these studies, declining alcohol consumption during quarantine was more common than increasing it. Although some of the observed differences across countries may be accounted by discrepancies in study designs (pre-post comparisons vs online surveys) and age of participants, they may also be influenced by the more social nature of drinking in Southern-Europe countries than in other cultures. In any case, to our knowledge, ours are the first results that are entirely based on an elderly population.

Results for changes in diet quality are difficult to compare because of differences in diet assessment across studies, with some showing the proportion of those who increased or decreased the consumption of certain products (11,15,56,57), others asking participants to report their self-perceived changes in quality (55,58), and only a few studies providing validated diet quality scores (17,18,59). Even for the latter, comparisons are limited due to the use of different scales; for instance, in a Spanish study, confinement was associated with a mean increase of 0.8 points in the MEDAS score (17), in one French study it was associated with a mean reduction of 0.32 points in the simplified PNNS-GS2 index (18), while in another study it was associated with equivalent increases or decreases in the Alternative Healthy Eating Index (59). Other factors compounding this comparison are the retrospective collection of pre-confinement information, and the lack of focus on older adults.

Our results were somewhat expected because, during the State of Alarm, both older adults (the segment of the population that is more used to daily shopping for foodstuffs in Spain(60)) and the whole the population, inevitably had to decrease the frequency of shopping of foods that are typically consumed fresh (i.e. fruits, vegetables or fish). In line with our results, two Spanish surveys indicated a reduction in the consumption of these fresh foods in younger adults during confinement (17,22). Despite this, we observed some compensations in diet quality in older adults thanks to a decrease in the intake of non-typical Mediterranean Diet foods (i.e. red meat, sweets and cakes), or an increase in the consumption of legumes, which are typically prepared at home.

Previous surveys described mental health during confinement as an important predictor of reductions in diet quality, a fact confirmed in our study, where those with higher follow-up GDS-10 scores showed a higher risk of diet quality decline (Relative Risk Ratio (RRR) per 1 point increase in the GDS-10: 1.25; 95%CI: 1.13-1.39). We also identified a number of baseline predictors of diet decline with confinement: social isolation (which may reflect not having someone to buy fresh foods for them), a lack of outdoor views (which is associated with lower income), the insufficiency of physical activity, or the suffering poor general health or hypertension.

Changes in weight occurring in short periods of time may be important if, as the literature on obesity suggests, they remain over time (61). Fortunately, though, most surveys either showed none or small changes in weight with confinement (14–16). In our study, despite mean declines in diet quality and physical activity, only specific subgroups of older adults (mainly hypertensives and those with a negative ageing experience) presented significant weight gains. Moreover, we observed that women, those with university studies, and those with short sleep and very long naps presented significant weight loss. The fact that weight loss was not associated with either baseline or follow-up (secondary analysis) overall health, psychological distress or depression symptoms, reduces the probability that it was involuntary and, thus, indicative of an increased risk of frailty (62). In any case, underlying changes in body composition (i.e. reduction in muscle mass and increase in fat mass) associated with the COVID-9 lockdown deserve further studies in the older population.

### What do other studies say about changes in physical activity and sedentary time, during the COVID-19 pandemic?

Surveys on non-institutionalized adults have uniformly shown declines in physical activity and increases in sedentary time after confinement (11,19,20,63–65), with unhealthier changes observed among those with higher activity and lower sedentariness at baseline (19,63,65), and among those who had follow-up symptoms of loneliness and worse mental health (12,19,63,65). In our study, we corroborated the association between previous levels of activity and sedentariness and changes in these behaviors with confinement, probably reflecting the regression to the mean phenomenon. Additionally, we first identified subpopulations of older adults who may be at increased risk of decreasing PA or increasing ST during quarantine. This information can help prioritize subgroups for health promotion or for being allowed to go outside, may future home confinements take place (i.e. the oldest old, those with worse housing conditions, or individuals with diabetes).

### What do other studies say about changes in sleep duration and sleep quality during the COVID-19 pandemic?

Worse sleep quality with confinement has been reported among the adult general population in different countries (11–13,65), and partly attributed to inactivity (12) and higher anxiety and mental distress during isolation (21). In our study, the association with mental health during quarantine was confirmed, with an increased risk of reducing sleep quality in individuals with higher GDS-10 scores (RRR: 1.25; 95%CI: 1.13-1.39; data not shown in tables). Moreover, our study identified for the first time predictors of these changes, showing that older women who live alone and feel lonely, those with worse diet quality, as well as those with worse overall health and higher pain are at increased risk of developing sleep problems with confinement.

### What do other studies say about changes in overall health and pain during the COVID-19 pandemic?

A mount of evidence supports a link between confinement and declines in mental health, both in middle-aged and older adults, and provides evidence that female gender (9) and being younger (66) are risk factors of these declines. However, we have only found one study evaluating the effect of confinement on chronic pain, this showing a mild improvement of migraine features during quarantine (67). Now we have identified subgroups of older adults who are more vulnerable to mental health declines with confinement, including those who feel lonelier, perform less physical activity, have mobility limitations, low MMSE scores or a negative ageing experience. Also, we found that individuals who lacked a garden/yard, with poor sleep quality, hypertension, cardiovascular disease, cancer, depression or mobility limitations were at increased risk of deteriorating physical function during confinement. Finally, we observed that women with short sleep, worse overall health and lower overall life satisfaction, osteomuscular disease and depression, as well as those who suffered from mobility limitations were at increased risk of pain worsening.

### Study limitations and strengths

Among the limitations of the present study is that not all cohorts had information in all the studied variables and a lack of data on the longer-term (July-December) effects of isolation measures. Moreover, the study cohort did not include institutionalized older adults, so results may not be generalizable to them. However, ours is one of the very few studies that have evaluated the effects of the pandemic in non-COVID-19-infected older adults. Also, unlike most of the reviewed web-based surveys, it relied on telephone interviews and used pre-pandemic in-home collected information on participants’ health behaviors and health status, reducing the risk of information bias (i.e. people not accurately reporting their past exposures or symptoms).

## Conclusion

The lockdown during the first wave of the COVID-19 in Spain, which was one of the most restrictive in Europe, only led to minor average changes in health behaviors among older adults. However, mental health was affected, particularly in those living alone, feeling lonelier, with mobility limitations and lower cognitive function. If another lockdown is imposed in this and future pandemics, public health programs should specially address the needs of older individuals of male sex, with greater social isolation, poor housing conditions and chronic morbidities, because of their greater vulnerability to the enacted movement restrictions.

## Data Availability

Data are available upon reasonable request to the authors.

## Declarations

### Ethics approval and consent to participate

All participants gave informed written consent and the different research protocols were approved by the Clinical Research Ethics Committee of the La Paz University Hospital in Madrid (*ENRICA*); the Clinical Research Ethics Review Committees of both Parc Sanitari Sant Joan de Déu and Hospital Universitario La Princesa (*ES*); the Clinical Research Ethics Committee of Toledo Hospital Complex (*TSHA*); and the Clinical Research Ethics Committee of Aragón (*EXERNET*)

### Consent for publication

NA

### Availability of data and materials

Data are available from the corresponding author on reasonable request

## Authors’ contributions

EGE, JMH, JLAM, LRM, IA, MM and FRA conceptualized the study. EGE, RO, IGV, JAC and AM conducted statistical analyses. EGE and FRA interpreted the results and drafted the initial manuscript. All authors reviewed the manuscript for important intellectual content and approved the final version as submitted.

## List of abbreviations

BMI: Body mass index
CAPI: Computer-Assisted Personal Interviews
COURAGE: Collaborative Research on Ageing in Europe
ES: Edad con Salud
ENRICA: Seniors-ENRICA-2
EXERNET: The elderly-Exernet multi-center study
MEDAS: Mediterranean diet adherence screener questionnaire
METs: Metabolic Equivalent Tasks
PASE: Physical Activity Scale for the Elderly
TSHA: Toledo Study for Healthy Ageing
WHODAS: WHO Disability Assessment Schedule 2.0

