## Supplemental Material for "Changes in health behaviors, mental and physical health among older adults under severe lockdown restrictions during the COVID-19 pandemic in Spain"

[1] Department of Preventive Medicine and Public Health. School of Medicine. Universidad Autónoma de Madrid, Madrid. Spain
[2] IdiPaz (Instituto de Investigación Sanitaria Hospital Universitario La Paz), Madrid. Spain
[3] CIBERESP (CIBER of Epidemiology and Public Health), Madrid, Spain.
[4] Parc Sanitari Sant Joan de Déu, Sant Boi de Llobregat, Spain.
[5] CIBERSAM (CIBER of Mental Health), Madrid, Spain.
[6] Foundation for Biomedical Research, Getafe University Hospital, Getafe, Spain.
[7] CIBERFES (CIBER of Frailty and Healthy Aging), Instituto de Salud Carlos III, Madrid, Spain.
[8] GENUD Toledo Research Group, Universidad de Castilla-La Mancha, Toledo, Spain.
[9] GENUD (Growth, Exercise, NUtrition and Development) research group, Instituto Agroalimentario de Aragón -IA2- (CITA-Universidad de Zaragoza)
[10] Department of Psychiatry. School of Medicine. Universidad Autónoma de Madrid, Spain.
[11] Department of Psychiatry, Hospital Universitario de La Princesa, Instituto de Investigación Sanitaria Princesa (IIS-Princesa), Madrid, Spain.
[12] CIBEROBN (CIBER of Obesity and Nutrition), Madrid, Spain.
[13] Faculty of Health and Sport Science, Huesca, and Department of Physiatry and Nursing, Zaragoza, University of Zaragoza, Spain.
[14] Department of Environmental Health, Harvard T.H. Chan School of Public Health. Boston, MA, USA.
[15] IMDEA-Food Institute, CEI UAM+CSIC, Madrid, Spain.
[16] Hospital Virgen del Valle, Complejo Hospitalario de Toledo, Toledo, Spain.
[17] AFYCAV (Physical Activity, Quality of Life and Health) research group, Faculty of Sport Sciences, University of Extremadura, Cáceres, Spain
[18] Department of Medicine. School of Medicine and Health Sciences. Universidad de Oviedo / ISPA, Oviedo Spain

**[*]Corresponding author:**
Dr. Esther García-Esquinas
Departamento Medicina Preventiva y Salud Pública
Universidad Autónoma de Madrid, Spain
Calle del Arzobispo Morcillo 4.
28029 Madrid, Spain
Phone: (+34) 91-497-27-61

**Abstract**

**Background:** We aimed to examine main changes in health behaviors, mental and physical health among older adults under severe lockdown restrictions during the COVID-19.

**Methods:** We used prospective data from 3041 participants in four cohorts of community-dwelling individuals aged ≥65 years in Spain. Data were obtained using validated questionnaires through a pre-pandemic face-to-face interview and a telephone interview conducted between weeks 7 to 15 after the beginning the COVID-19 lockdown. Lineal or multinomial, as appropriate, regression models with adjustment for the main confounders were used to assess changes in the outcome variables from the pre-pandemic to the confinement period, and to identify their associated factors.

**Results:** On average, the confinement was not associated with a deterioration in lifestyle risk factors (smoking, alcohol intake, diet or weight), except for a decreased physical activity and increased sedentary time, which reversed with the end of confinement. However, chronic pain worsened, and moderate declines in mental health, that did not seem to reverse after restrictions were lifted, were observed. Several subgroups of individuals were at increased risk of developing unhealthier lifestyles or mental health decline with confinement: (i)-males (for physical activity and sedentariness), (ii)-those with greater social isolation (for diet, physical activity, mental health), (iii)-feelings of loneliness (for diet, sleep quality, mental health), (iv)-poor housing conditions (for diet, physical activity, TV viewing time), (v)-unhealthy sleep duration (for physical activity and sedentariness), and (vi-worse overall health or chronic morbidities (for physical activity, screen time, mental health). On the other hand, previously having a greater adherence to the Mediterranean diet and doing more physical activity protected older adults from developing unhealthier lifestyles with confinement.

**Conclusions:** The lockdown during the first wave of the COVID-19 in Spain, which was one of the most restrictive in Europe, only led to minor average changes in health behaviors among older adults. However, mental health was moderately affected. If another lockdown were imposed on this or future pandemics, public health programs should specially address the needs of older individuals with male sex, greater social isolation, poor housing conditions and chronic morbidities, because of their greater vulnerability to the enacted movement restrictions

**Supplementary table 1**: Prevalence or mean (SD) for the main sociodemographic, lifestyle and health-related characteristics of older adults at the COVID-19 pre-confinement period (t0) and the State of Alarm (t1)†

| | *ENRICA* (n=1323) | | *ES* (n=464) | | *TSHA* (n=829) | | **Exernet** (n=425) | |
|---|---|---|---|---|---|---|---|---|
| | **t0** | **t1** | **t0** | **t1** | **t0** | **t1** | **t0** | **t1** |
| **Socio-demographic variables** | | | | | | | | |
| **Age**; $\bar{x}$ (SD) | 73.7(4.3) | 75.4(4.3) | 69.9(8.0) | 69.9(8.0) | 76.8(5.5) | 81.5(5.6) | 77.8(4.5) | 81.3(4.5) |
| **Male**; % | 50.5 | | 42.9 | | 39.8 | | 21.2 | |
| **University**; % | 22.3 | | 16.4 | | 8.2 | | 8.0 | |
| **Civil status**; % | | | | | | | | |
| Married | 65.2 | | 64.2 | | 68.0 | | 57.4 | |
| Single | 7.0 | | 4.1 | | 4.2 | | 4.0 | |
| Divorced | 5.9 | | 10.3 | | 2.1 | | 2.6 | |
| Widowed | 21.9 | | 21.3 | | 25.7 | | 35.9 | |
| **Living alone**; % | 23.6 | 24.1 | 21.6 | 18.5 | 21.2 | 25.2 | 34.1 | 35.8 |
| **No daily contact** with family/friends; % | 12.9 | 7.3 | 47.9 | 20.8 | 26.5 | 10.6 | | |
| **Feeling lonely (1-5 scale)**; $\bar{x}$ (SD) | 1.6 (0.9) | 1.4 (0.9) | 1.5 (1.1) | | | | | |
| **Housing conditions** | | | | | | | | |
| **No outdoor views**; % | 8.2 | | 9.1 | | 4.3 | | 9.9 | |
| **No terrace/balcony**; % | 25.7 | | 32.8 | | 37.0 | | 24.0 | |
| **No private garden/yard**; % | 86.6 | | 72.8 | | 41.9 | | 72.2 | |
| **Lack of internet**; % | 20.5 | | 23.1 | | 54.9 | | 63.3 | |
| **Too much noise**; % | 2.3 | | 4.7 | | 3.1 | | 3.3 | |
| **Size of house (m$^2$)**; $\bar{x}$ | 95.6 | | | | | | | |
| **Lifestyle-behaviours** | | | | | | | | |
| **Smoking**; % | | | | | | | | |
| Never smokers | 51.1 | 52.1 | 54.6 | 50.0 | 69.9 | 71.7 | 79.5 | 89.9 |
| Former smokers | 39.4 | 40.9 | 28.9 | 34.5 | 23.1 | 24.5 | 16.7 | 9.4 |
| Current smoker | 8.9 | 7.0 | 16.4 | 15.5 | 6.8 | 3.7 | 1.7 | 0.7 |
| **Exposure to SHS**; % | 4.3 | 9.9 | | | | | | |
| **Alcohol intake**; % | | | | | | | | |
| Not drinker | 42.6 | 46.0 | 54.1 | 52.4 | 62.3 | 66.1 | | |
| Drinker, not daily | 28.0 | 19.1 | 29.1 | 26.5 | 10.1 | 10.6 | | |
| Drinker, daily/almost daily | 29.4 | 35.0 | 16.8 | 21.1 | 27.6 | 23.3 | | |
| **MEDAS index**; $\bar{x}$ (SD) | 7.2 (1.7) | 7.4 (1.7) | 8.5 (1.8) | 7.5 (1.7) | 7.4 (1.7) | 6.6 (1.8) | 9.0 (2.0) | 7.4 (1.6) |
| **Eating at a regular time; %** | | 94.7 | | - | | 96.8 | | 96.5 |
| **Eating more ultra-processed, %** | | 2.3 | | 5.3 | | 0.6 | | 0.2 |
| **Eating snacks** | 45.6 | 37.3 | | | | | | |
| **Eating to calm anxiety** | 13.2 | 8.1 | | | | | | |
| **rPA (Mets-h/week)**; $\bar{x}$ (SD) | 53.2 (24.9) | 43.1 (27.3) | 23.5 (37.5) | ‡ | | | 81.8 (40.0) | 36.0 (27.1) |
| **hPA Mets-h/week)**; $\bar{x}$ (SD) | 24.8 (19.6) | 23.6 (21.3) | | | | | 44.6 (34.4) | 22.0 (22.5) |
| **PASE score**; $\bar{x}$ (SD) | | | | | 87.8 (44.0) | 69.7 (52.6) | | 73.8 (60.0) |
| **BMI** | | | | | | | | |
| Normoweight | 29.0 | | 27.8 | | 14.8 | | 19.5 | |
| Overweight | 48.9 | | 43.1 | | 42.0 | | 46.1 | |
| Obese | 23.1 | | 29.1 | | 43.2 | | 34.4 | |
| **Weight;**[§] $\bar{x}$ (SD) | 72.2 (12.7) | 71.8 (12.1) | 73.8 (13.9) | 73.6 (13.3) | 73.4 (13.1) | 71.7 (13.0) | 69.3 (11.1) | 67.9 (11.0) |

| | *ENRICA* (n=1323) | | *ES* (n=464) | | *TSHA* (n=829) | | **Exernet** (n=425) | |
|---|---|---|---|---|---|---|---|---|
| | **t0** | **t1** | **t0** | **t1** | **t0** | **t1** | **t0** | **t1** |
| **Total ST (h/d)**; $\bar{x}$ (SD) | 6.1 (2.7) | 7.2 (3.2) | 4.8 (2.5) | ‡ | | 6.7 (3.2) | 4.3 (2.1) | 6.8 (3.3) |
| **TV viewing time (h/d);** $\bar{x}$ (SD) | 3.5 (1.8) | 4.0 (2.2) | | | | | | |
| **Other screen time** (min/day); $\bar{x}$ (SD) | 36.5 (64.8) | 82.0 (96.4) | | | | | | |
| **Reading** (min/day)**; $\bar{x}$ (SD)** | 51.8 (58.5) | 68.2 (74.7) | | | | | | |
| **Listening to music (min/day); $\bar{x}$ (SD)** | 11.3 (33.6) | 19.2 (49.6) | | | | | | |
| **Sleep characteristics** | | | | | | | | |
| **Hours of night-time sleep**; % | | | | | | | | |
| Normal sleep | 79.2 | 73.0 | 77.2 | 75.0 | 62.6 | 62.7 | 66.6 | 67.1 |
| Short sleep (≤6 h) | 16.3 | 18.2 | 11.9 | 16.4 | 18.1 | 18.5 | 25.4 | 14.4 |
| Long sleep (≥9 h) | 4.5 | 8.8 | 11.0 | 8.6 | 19.3 | 18.8 | 8.0 | 18.5 |
| **Hours of day-time sleep**; % | | | | | | | | |
| None | 394 | 717 | | | | | 205 | 236 |
| Short nap (≤30 min) | 623 | 556 | | | | | 125 | 180 |
| Long nap (30-60 min) | 226 | 41 | | | | | 62 | 5 |
| Very long nap (≥60min) | 71 | 0 | | | | | 29 | 0 |
| **Overall sleep quality**; % | | | | | | | | |
| Very good | 65.1 | 62.6 | 47.4 | 72.6 | | | 46.2 | 64.6 |
| Good | 10.8 | 11.2 | 41.8 | 11.4 | | | 23.0 | 6.1 |
| Fair | 21.8 | 22.0 | 9.9 | 13.7 | | | 23.2 | 25.1 |
| Poor/very poor | 2.3 | 4.2 | 0.9 | 2.3 | | | 7.7 | 4.3 |
| **Poor sleep quality score**;‖ $\bar{x}$ (SD) | 1.7 (1.5) | 1.8 (1.5) | | | | | | |
| **Health-related variables** | | | | | | | | |
| **Overall health**; $\bar{x}$ (SD) | | | | | | | | |
| SF-12, PCS | 46.0 (11.0) | 50.2 (8.6) | | | | | | |
| SF-12, MCS | 54.1 (9.6) | 53.2 (8.5) | | | | | | |
| WHODAS-12 | | | 9.4 (15.5) | 11.9 (17.1) | | | | |
| EQ-5D | | | | | 0.9 (0.2) | | 0.8 (0.2) | |
| **Pain scale***; $\bar{x}$ (SD) | 0.9 (2.2) | 1.4 (2.2) | | | | | | |
| **Mobility limitations;** % | 36.0 | | | | | | | |
| **Negative ageing experience scale*;** $\bar{x}$ (SD) | 2.5 (1.0) | | | | | | | |
| **Cantril ladder***; $\bar{x}$ (SD) | 8.1 (1.8) | | | | | | | |
| **Low MMSE scores (<23)**; % | 3.4 | | | | | | | |
| **Week of state of alarm**; † range | 7-11 | | 10-15 | | 8-11 | | 7-11 | |

ENRICA: Seniors-ENRICA-2. ES: Edad con Salud. TSH: Toledo Study for Healthy Ageing.

$\bar{x}$: Mean; SD: Standard deviation; MEDAS: Mediterranean Diet Assessment Score; rPA: Recreational physical activity; hPA: Household physical activity; BMI: Body Mass Index; ST: Sedentary time; PCS: Physical Component Score of the SF-12; MCS: Mental Component Score of the SF-12

†The Spanish Government declared the state of alarm on March 14th, meaning general lockdown for the entire country from March 15th. From May 4th (week 8) adults were allowed out to walk and exercise a maximum of 1 hour/dat during set time slots. The de-escalation phase was gradual during the following weeks, until the country started its "New Normality" phase on June 22nd (week 14)

‡ Differences between questionnaires did not allow for pre-post comparisons

§ Information at follow-up was only available in the subsample of participants that had weight themselves on the week of the telephone interview (n=1566)

‖ The score ranged 0 (best) to 7 (worse sleep). Participants who answered "sometimes" or "almost always" to four indicators of poor nighttime sleep (i.e. difficulty falling asleep, awakening during nighttime, early awakening with difficulty getting back to sleep, use of sleeping pills), two indicators of drowsiness (being so sleepy at daytime as to need a nap, not feeling rested in the morning), or with a score >10 in the Epworth Sleepiness Scale, received 1 point; their counterparts received 0 points

¶ Higher scores in the MCS and PCS of the SF-12 as well as on the EQ-5D, and lower scores in the WHODAS-12 are indicative of better health.

*The pain scale ranged 0 (best) to 7 (worse pain) and included information on frequency, intensity and number of pain sites. Individuals with sporadic (<2 time/day) and frequent (≥2 times/week ) pain were assigned a score of 1 and 2, respectively; those with light, moderate and high intensity pain were given a score of 1,2 and 3; and those with 1-2 and ≥3 locations of pain a score of 1 and 2, respectively. The ageing experience scale ranged from 1 (very positive experience) to 5 (very negative experience). The Cantril ladder ranged from 1 (less satisfied) to 10 (more satisfied).

**Supplementary table 2.** Prospective association between baseline participant characteristics and changes in frequency of tobacco (n=194: 93 from *Seniors ENRICA-2*, 67 from *Edad con Salud*, 31 from *TSH*A, and 3 from *Exernet*) and alcohol (n=2616: 1323 from *Seniors ENRICA-2*, 464 from *Edad con Salud*, and 829 from *TSH*A) consumption between the COVID-19 pre-confinement period and the State of Alarm

| | **Changes in frequency of tobacco consumption** | | | | **Changes in frequency of alcohol consumption** | | | | |
|---|---|---|---|---|---|---|---|---|---|
| | **n** | **Same frequency** | **Increased frequency** | **Decreased frequency** | **n** | **Same frequency** | **Increased frequency** | **Decreased frequency** | **Maintained non-drinking status** |
| | | RRR(95%CI) n=99 | RRR (95%CI) n=42 | RRR (95%CI) n=53 | | RRR(95%CI) n=731 | RRR (95%CI) n=400 | RRR (95%CI) n=447 | RRR (95%CI) n=1038 |
| **Socio-demographic characteristics** | | | | | | | | | |
| **Age, yr** | 194 | Ref. | 1.02 (0.92;1.13) | 1.02 (0.92;1.13) | 2616 | Ref. | **1.03 (1.00;1.05)** | **1.03 (1.00;1.06)** | **1.03 (1.01;1.06)** |
| **Sex** | | | | | | | | | |
| Male | 114 | Ref. | Ref. | Ref. | 1197 | Ref. | Ref. | Ref. | Ref. |
| Female | 80 | Ref. | 1.27 (0.45;3.58) | 0.80 (0.31;2.09) | 1419 | Ref. | **1.55 (1.14;2.11)** | **1.95 (1.45;2.71)** | **4.65 (3.58;6.04)** |
| **Education** | | | | | | | | | |
| Primary or less | 102 | Ref. | Ref. | Ref. | 1694 | Ref. | Ref. | Ref. | Ref. |
| Secondary | 49 | Ref. | 2.09 (0.67;6.57) | 0.74 (0.26;2.16) | 483 | Ref. | 1.03 (0.74;1.43) | 1.01 (0.73;1.39) | **0.71 (0.53;0.95)** |
| University | 43 | Ref. | 1.94 (0.58;6.54) | 0.91 (0.29;2.86) | 439 | Ref. | 0.89 (0.62;1.26) | 1.03 (0.74;1.43) | **0.62 (0.45;0.85)** * |
| **Civil status** | | | | | | | | | |
| Married | 122 | Ref. | Ref. | Ref. | 1725 | Ref. | Ref. | Ref. | Ref |
| Never married | 11 | Ref. | 0.46 (0.03;7.32) | 3.87 (0.44;34.3) | 146 | Ref. | 0.67 (0.33;1.36) | 1.07 (0.60;1.92) | 1.05 (0.62;1.79) |
| Divorced | 20 | Ref. | 0.49 (0.07;3.35) | 0.26 (0.04;1.90) | 143 | Ref. | 0.73 (0.37;1.43) | 1.10 (0.60;2.00) | 1.22 (0.72;2.06) |
| Widowed | 41 | Ref. | 0.74 (0.15;3.51) | **0.19 (0.04;0.89)** | 602 | Ref. | 1.25 (0.78;1.99) | 0.90 (0.56;1.42) | 1.33 (0.90;1.96) |
| **Living alone** | | | | | | | | | |
| No | 151 | Ref. | Ref. | Ref. | 2028 | Ref. | | Ref. | Ref. |
| Yes | 43 | Ref. | 1.74 (0.41;7.35) | 1.33 (0.26;6.71) | 588 | Ref. | 1.06 (0.65;1.71) | 1.30 (0.82;2.07) | 0.06 (0.65;1.43) |
| **Daily contact family/friends** | | | | | | | | | |
| Yes | 137 | Ref. | Ref. | Ref. | 1989 | Ref. | Ref. | Ref | Ref. |
| No | 54 | Ref. | 1.01 (0.34;2.93) | 1.83 (0.71;4.72) | 623 | Ref. | 0.88 (0.63;1.23) | 0.86 (0.63;1.19) | 1.17 (0.89; 1.54) |
| **Feeling lonely (1-5 scale)†** | 159 | Ref. | 1.05 (0.65;1.69) | 0.96 (0.57;1.63) | 1776 | Ref. | 1.15 (0.98;1.36) | 1.03 (0.87;1.23) | 1.10 (0.96;1.28) |
| **Housing conditions** | | | | | | | | | |
| **Lack of outdoor views** | | | | | | | | | |
| No | 186 | Ref. | Ref. | Ref. | 186 | Ref. | Ref. | Ref. | Ref. |
| Yes | 8 | Ref. | 0.74 (0.06;9.77) | 3.21 (0.42;24.3) | 2430 | Ref. | **1.75 (1.03;3.10)** | **2.23 (1.35;3.68)** | **2.30 (1.44;3.66)** |
| **Lack of terrace/balcony** | | | | | | | | | |
| No | 148 | Ref. | Ref. | Ref. | 1817 | Ref. | Ref. | Ref | Ref. |
| Yes | 46 | Ref. | **0.34 (0.10;1.00)** | 0.95 (0.32;2.77) | 799 | Ref. | **0.69 (0.51;0.93)** | 1.18 (0.90;1.55) | 1.25 (0.99;1.59) |

| | Changes in frequency of tobacco consumption | | | | Changes in frequency of alcohol consumption | | | | |
|---|---|---|---|---|---|---|---|---|---|
| | n | Same frequency | Increased frequency | Decreased frequency | n | Same frequency | Increased frequency | Decreased frequency | Maintained non-drinking status |
| | | RRR(95%CI) n=99 | RRR (95%CI) n=42 | RRR (95%CI) n=53 | | RRR(95%CI) n=731 | RRR (95%CI) n=400 | RRR (95%CI) n=447 | RRR (95%CI) n=1038 |
| **Lack of garden/yard** | | | | | | | | | |
| No | 39 | Ref. | Ref. | Ref. | 789 | Ref. | Ref. | Ref. | Ref. |
| Yes | 144 | Ref. | 2.76 (0.77;9.88) | 0.81 (0.29;2.28) | 1830 | Ref. | 0.90 (0.66;1.23) | 1.14 (0.84;1.55) | **1.33 (1.02;1.74)** |
| **Internet access** | | | | | | | | | |
| Yes | 151 | Ref. | Ref. | Ref. | 1783 | Ref. | Ref. | Ref. | Ref. |
| No | 43 | Ref. | 1.92 (0.56;6.66) | **0.31 (0.00;1.04)** | 833 | Ref. | 1.21 (0.88;1.68) | 1.10 (0.80;1.51) | **1.61 (1.23;2.11)** |
| **Too much noise** | | | | | | | | | |
| No | 188 | Ref. | Ref. | Ref. | 2537 | Ref. | Ref. | Ref. | Ref. |
| Yes | 6 | Ref. | 1.45 (0.09;23.6) | 1.97 (0.22;17.4) | 79 | Ref. | 0.72 (0.32;1.64) | 1.13 (0.56;2.27) | 0.89 (0.47;1.66) |
| **Lifestyle-behaviours** | | | | | | | | | |
| **Smoking** | | | | | | | | | |
| Never smokers | - | Ref. | - | - | 1507 | Ref. | Ref. | Ref. | Ref. |
| Former smokers | - | Ref. | - | - | 853 | Ref. | **0.59 (0.44;0.80)** | 0.94 (0.71;1.25) | **0.59 (0.45;0.76)** |
| Smoker | - | Ref. | - | - | 247 | Ref. | 0.88 (0.57;1.35) | 1.14 (0.75;1.73) | **0.63 (0.43;0.94)** |
| **MEDAS** | 186 | Ref. | 1.17 (0.54;6.92) | 1.11 (0.87;1.40) | 2616 | Ref. | **0.93 (0.86;0.99**) | 0.96 (0.89;1.03) | **0.92 (0.86;0.98)** |
| **PA** (cohort-specific quartiles) | | | | | | | | | |
| 1st quartile (less activity) | 56 | Ref. | Ref. | Ref. | 676 | Ref. | Ref. | Ref. | Ref. |
| 2nd quartile | 40 | Ref. | 0.40 (0.11;1.45) | 1.08 (0.31;3.71) | 667 | Ref. | 1.02 (0.72;1.44) | 1.21 (0.85;1.72) | 1.00 (0.73;1.37) |
| 3rd quartile | 49 | Ref. | 0.54 (0.16;1.85) | 1.43 (0.46;4.43) | 665 | Ref. | 0.79 (0.55;1.14) | 1.08 (0.76;1.54) | 0.86 (0.62;1.18) |
| 4th quartile | 49 | Ref. | 0.99 (0.30;3.28) | 1.61 (0.50;5.20) | 608 | Ref. | 0.71 (0.49;1.04) | 0.97 (0.67;1.40) | 1.03 (0.74;1.41) |
| **BMI** | | | | | | | | | |
| Normoweight | 73 | Ref. | Ref. | Ref. | 601 | Ref. | Ref. | Ref. | Ref. |
| Overweight | 83 | Ref. | 0.82 (0.30;2.24) | **2.60 (1.00;6.77)** | 1216 | Ref. | 1.19 (0.86;1.65) | 1.05 (0.77;1.42) | 1.07 (0.82;1.42) |
| Obese | 38 | Ref. | 0.73 (0.20;2.59) | **2.97** (0.97;9.67) | 799 | Ref. | 1.26 (0.87;1.84) | 1.15 (0.81;1.65) | 1.24 (0.90;1.70) |
| **ST** (cohort-specific quartiles)† | | | | | | | | | |
| 1st quartile (less time) | 47 | Ref. | Ref. | Ref. | 504 | Ref. | Ref. | Ref. | Ref |
| 2nd quartile | 21 | Ref. | **8.21 (1.54;43.7)** | 0.24 (0.02;2.54) | 407 | Ref. | 0.89 (0.60;1.34) | 1.27 (0.84;1.92) | 0.87 (0.61;1.25) |
| 3rd quartile | 48 | Ref. | 2.21 (0.57;8.48) | 1.37 (0.38;4.97) | 444 | Ref. | **0.61 (0.40;0.92)** | 1.00 (0.66;1.51) | 0.84 (0.59;1.20) |
| 4th quartile | 47 | Ref. | 1.88 (0.45;7.82) | 3.00 (0.72;12.3) | 432 | Ref. | 0.98 (0.66;1.46) | 1.32 (0.87;2.00) | 0.79 (0.54;1.14) |
| **Sleep characteristics** | | | | | | | | | |
| **Hours of night-time sleep** | | | | | | | | | |
| Normal sleep | 144 | Ref. | Ref. | Ref. | 1925 | Ref. | Ref. | Ref. | Ref. |
| Short sleep (≤6 h) | 28 | Ref. | 1.93 (0.54;6.92) | 0.39 (0.10;1.53) | 421 | Ref. | 1.19 (0.84;1.68) | 0.99 (0.70;1.41) | 1.11 (0.47;1.66) |
| Long sleep (≥9 h) | 22 | Ref. | 1.14 (0.26;4.97) | 0.86 (0.21;3.48) | 270 | Ref. | 1.42 (0.88;2.29) | 1.39 (0.88;2.18) | **1.77 (1.23;2.11)** |

| | Changes in frequency of tobacco consumption | | | | Changes in frequency of alcohol consumption | | | | |
|---|---|---|---|---|---|---|---|---|---|
| | n | Same frequency | Increased frequency | Decreased frequency | n | Same frequency | Increased frequency | Decreased frequency | Maintained non-drinking status |
| | | RRR(95%CI) n=99 | RRR (95%CI) n=42 | RRR (95%CI) n=53 | | RRR(95%CI) n=731 | RRR (95%CI) n=400 | RRR (95%CI) n=447 | RRR (95%CI) n=1038 |
| **Hours of day-time sleep**‡ | | | | | | | | | |
| None | 27 | Ref. | Ref. | Ref. | 395 | Ref. | Ref. | Ref. | Ref. |
| Short nap (≤30 min) | 40 | Ref. | 0.18 (0.02;1.44) | 1.47 (0.16;13.4) | 625 | Ref. | 0.88 (0.59;1.32) | 0.90 (0.61;1.33) | 0.81 (0.57;1.16) |
| Long nap (30-60 min) | 18 | Ref. | 0.32 (0.03;3.53) | 1.53 (0.12;19.1) | 229 | Ref. | 1.18 (0.71;1.93) | 0.69 (0.41;1.18) | 0.79 (0.49;1.26) |
| Very long nap (≥60min) | 11 | Ref. | 0.29 (0.02;4.75) | 0.99 (0.06;15.3) | 71 | Ref. | **0.39 (0.16;0.96)** | 1.04 (0.50;2.16) | 0.56 (0.26;1.19) |
| **Overall sleep quality**† | | | | | | | | | |
| Very good | 100 | Ref. | 0.73 (0.16;3.40) | 0.64 (0.19;2.22) | 337 | Ref. | 0.96 (0.64;1.45) | 1.07 (0.71;1.61) | 0.96 (0.64;1.45) |
| Good | 35 | Ref. | Ref. | Ref, | 1081 | Ref. | Ref. | Ref. | Ref. |
| Fair | 280 | Ref. | **6.16 (1.28;29.6)** | 0.66 (0.11;4.09) | 335 | Ref. | **0.65 (0.43;0.98)** | 0.78 (0.51;1.18) | **0.64 (0.43;0.98)** |
| Poor/very poor | | Ref. | - | - | 34 | Ref. | 1.24 (0.34;4.48) | **3.73 (1.15;12.1)** | 1.24 (0.34;4.48) |
| **Health variables** | | | | | | | | | |
| **Overall health** | | | | | | | | | |
| 1st quartile (worst) | 33 | Ref. | Ref. | Ref. | 512 | Ref. | Ref. | Ref. | Ref. |
| 2nd quartile | 51 | Ref. | 3.71 (0.69;20.0) | 1.23 (0.30;5.01) | 486 | Ref. | 1.09 (0.71;1.69) | 1.09 (0.70;1.71) | 1.05 (0.73;1.53) |
| 3rd quartile | 27 | Ref. | 1.53 (0.21;11.1) | 0.52 (0.10;2.81) | 523 | Ref. | 0.90 (0.58;1.38) | 1.05 (0.69;1.63) | **0.61 (0.42;0.89)** |
| 4th quartile (best) | 83 | Ref. | 1.60 (0.29;8.99) | 0.30 (0.07;1.25) | 1095 | Ref. | 0.82 (0.53;1.27) | 1.06 (0.69;1.64) | **0.51 (0.42;0.88)** * |
| **Pain scale**‡ | 77 | Ref. | 1.10 (0.56;2.15) | 0.67 (0.32;1.41) | 1049 | Ref. | 0.98 (0.88;1.09) | 0.95 (0.85;1.05) | 0.98 (0.89;1.08) |
| **Chronic morbidities** | | | | | | | | | |
| Diabetes | 28 | Ref. | 1.70 (0.51;5.70) | 0.40 (0.11;1.52) | 471 | Ref. | 0.95 (0.67;1.34) | 0.90 (0.64;1.27) | 1.30 (0.98;1.74) |
| Hypertension | 91 | Ref. | 0.51 (0.21;1.25) | 0.72 (0.32;1.64) | 1480 | Ref. | 1.28 (0.98;1.67) | 1.11 (0.86;1.44) | 1.02 (0.81;1.29) |
| CVD | 22 | Ref. | 5.23 (0.70;39.3) | 1.88 (0.30;11.9) | 199 | Ref. | 0.80 (0.47;1.34) | 1.10 (0.68;1.78) | 0.92 (0.60;1.41) |
| Cancer | 15 | Ref. | 1.40 (0.07;2.38) | **0.05 (0.00;0.46)** | 188 | Ref. | 1.03 (0.64;1.68) | 0.89 (0.55;1.44) | 1.02 (0.67; 1.55) |
| Osteomuscular disease | 67 | Ref. | 1.03 (0.37;2.88) | **0.**39 (0.15;1.06) | 1041 | Ref. | 0.89 (0.67;1.17) | 0.86 (0.66;1.13) | 0.79 (0.62;1.02) |
| Depression | 26 | Ref. | 0.78 (0.17;3.63) | 1.00 (0.23;4.32) | 311 | Ref. | 1.20 (0.75;1.91) | 1.15 (0.73;1.81) | 1.44 (0.97;2.11) |
| **Mobility limitations**‡ | 29 | Ref. | 3.57 (0.41;31.3) | 0.29 (0.03;2.70) | 476 | Ref. | 0.78 (0.50;1.22) | 0.95 (0.62;1.47) | 1.04 (0.71;1.54) |
| **Negative ageing experience scale** ‡ | 92 | Ref. | 0.91 (0.44;1.88) | 0.52 (0.24;1.12) | 1313 | Ref. | 0.99 (0.83;1.19) | 1.09 (0.91;1.30) | 0.97 (0.82;1.15) |
| **Cantril ladder**§ | 158 | Ref. | 1.28 (0.96;1.70) | 1.08 (0.82;1.43) | 1770 | Ref. | 1.03 (0.94;1.12) | 0.96 (0.88;1.04) | 1.02 (0.94;1.10) |
| **Low MMSE scores (<23)**§ | 6 | Ref. | 0.14 (0.00;3.29) | 0.74 (0.04;14.6) | 77 | Ref. | 1.07 (0.41;3.81) | 1.99 (0.80;4.79) | 1.68 (0.78;3.63) |
| **Years since baseline exam** | 194 | Ref. | 2.20 (0.60;8.03) | 3.31 (0.97;11.3) | 2616 | Ref. | 0.99 (0.75;1.31) | 0.93 (0.72;1.21) | 1.06 (0.84;1.34) |
| **Week of the state of alarm** | 194 | Ref. | 0.92 (0.60;1.41) | 0.97 (0.63;1.49) | 2616 | Ref. | 0.96 (0.84;1.10) | 1.01 (0.89;1.15) | 0.99 (0.88;1.11) |

OR: Odds Ratio. 95%CI: 95% Confidence interval.; MEDAS: Mediterranean Diet Assessment Score; PA: Physical activity; BMI: Body Mass index; ST: Sedentary time; MMSE: Mini Mental State Examination

Note: Figures highlighted in bold show a statistically significant association.

† Information not available in the *TSHA* cohort; ‡ Information only available in the *Seniors ENRICA-2* cohort; § Information only available in the *Seniors ENRICA-2* and *Edad con Salud* cohorts

All models were adjusted for baseline age, sex (men or women), educational level (primary or less, secondary, or university), civil status (married, widowed, never married, divorced), smoking status (never, former, or current), adherence to the Mediterranean diet (MEDAS score), body mass index (normoweight, overweight, or obese), physical activity (quartiles), hours of sleep at night (normal, short sleep, long sleep), chronic comorbidities (hypertension, diabetes, cardiovascular disease, cancer, osteo-muscular disease and depression), overall health (SF-12, WHODAS-12 o EQ5D), time since last follow-up visit, cohort of study and week of the state of alarm.
* p-value for trend <0.05

.

**Supplementary Table 3.** Prospective association between participant characteristics at baseline and changes in adherence to the MEDAS score between the COVID-19 pre-confinement period and the State of Alarm (n=3041: 1323 from *Seniors ENRICA-2*, 464 from *Edad con Salud*, 829 from *TSHA*, and 425 from *EXERNET*).

| | | Changes in MEDAS | | | |
|---|---|---|---|---|---|
| | **n** | **No changes** | **Decreased ≥1 point** | **Increased ≥1 point** | **Mean changes** |
| | | | OR (95%CI) | OR (95%CI) | $\bar{x}$ (95%CI) |
| | | n=162 | n=949 | n=470 | n=3045 |
| **Socio-demographic characteristics** | | | | | |
| **Age, yr** | 3041 | Ref. | **1.02 (1.00; 1.04)** | 0.98 (0.96; 1.01) | **-0.02 (-0.03; -0.01)** |
| **Sex** | | | | | |
| Male | 1287 | Ref. | 1.00 | Ref. | Ref. |
| Female | 1754 | Ref. | 0.93 (0.73; 1.19) | 0.89 (0.68; 1.18) | 0.11 (-0.03; 0.25) |
| **Education** | | | | | |
| Primary or less | 2040 | Ref. | Ref. | Ref. | Ref. |
| Secondary | 528 | Ref. | 0.94 (0.72; 1.22) | 1.02 (0.75; 1.38) | 0.09 (-0.06; 0.25) |
| University | 473 | Ref. | **0.73 (0.54; 0.99)** * | 0.89 (0.64; 1.24) | **0.13** (-0.04; 0.30) |
| **Civil status** | | | | | |
| Married | 1969 | Ref. | 1.00 | 1.00 | Ref. |
| Never married | 163 | Ref. | 0.99 (0.61; 1.61) | 0.76 (0.42; 1.37) | -0.02 (-0.30; 0.27) |
| Divorced | 154 | Ref. | 1.11 (0.67; 1.83) | 0.98 (0.55; 1.73) | -0.15 (-0.44; 0.14) |
| Widowed | 755 | Ref. | 1.13 (0.81; 1.56) | 1.05 (0.72; 1.55) | **-0.20 (-0.39; -0.01)** |
| **Living alone** | | | | | |
| No | 2308 | Ref. | Ref. | Ref. | Ref. |
| Yes | 733 | Ref. | 1.07 (0.77; 1.49) | 0.91 (0.61; 1.35) | 0.04 (-0.15; 0.23) |
| **Daily contact with family/friends other than cohabitants**† | | | | | |
| Yes | 1989 | Ref. | Ref. | Ref. | Ref. |
| No | 623 | Ref. | 1.26 (0.97; 1.63) | 0.79 (0.57; 1.10) | **-0.26 (-0.42; -0.10)** |
| **Feeling lonely (1-5 scale)**‡ | 1776 | Ref. | 1.12 (0.97; 1.29) | 0.94 (0.80;1.10) | **-0.08 (-0.16; -0.01)** |
| **Housing conditions** | | | | | |
| **Lack of outdoor views** | | | | | |
| No | 2816 | Ref. | Ref. | Ref. | Ref. |
| Yes | 228 | Ref. | **1.53 (1.05; 2.21)** | 0.90 (0.58; 1.40) | **-0.29 (-0.50; -0.07)** |
| **Lack of terrace/balcony** | | | | | |
| No | 2140 | Ref. | Ref. | Ref. | Ref. |
| Yes | 904 | Ref. | 1.10 (0.88; 1.36) | 1.09 (0.85; 1.41) | -0.08 (-0.21; 0.04) |
| **Lack of private garden/yard** | | | | | |
| No | 907 | Ref. | Ref. | Ref. | **Ref.** |
| Yes | 2137 | Ref. | 1.09 (0.87; 1.37) | 0.85 (0.64; 1.12) | -0.05 (-0.19; 0.08) |

| | n | No changes | Changes in MEDAS Decreased ≥1 point | | Increased ≥1 point | Mean changes | |
|---|---|---|---|---|---|---|---|
| | | | OR (95%CI) | | OR (95%CI) | x̄ (95%CI) | |
| | | n=162 | n=949 | | n=470 | n=3045 | |
| **Internet access** | | | | | | | |
| Yes | 1939 | Ref. | Ref. | | **Ref.** | **Ref.** | |
| No | 1105 | Ref. | 1.10 (0.88; 1.38) | | **0.74 (0.56; 0.98)** | **-0.16 (-0.29; -0.03)** | |
| **Too much noise** | | | | | | | |
| No | 2951 | Ref. | Ref | | Ref. | **Ref.** | |
| Yes | 93 | Ref. | 1.52 (0.85; 2.68) | | 0.99 (0.55; 1.87) | -0.23 (-0.56; 0.09) | |
| **Lifestyle-behaviours** | | | | | | | |
| **Smoking** | | | | | | | |
| Never smokers | 1863 | Ref. | Ref. | | Ref. | Ref. | |
| Former smokers | 924 | Ref. | 0.88 (0.68; 1.14) | | 0.99 (0.75; 1.29) | 0.09 (-0.05; 0.24) | |
| Smoker | 254 | Ref. | 1.35 (0.92; 1.99) | | 1.05 (0.71; 1.57) | -0.14 (-0.37; 0.07) | |
| **Alcohol intake**† | | | | | | | |
| Not drinker | 1330 | Ref. | Ref. | | Ref. | Ref. | |
| Drinker, not daily | 590 | Ref. | 0.77 (0.58; 1.03) | | 0.85 (0.62; 1.16) | 0.10 (-0.06; 0.26) | |
| Drinker, daily or almost daily | 696 | Ref. | **0.63 (0.47; 0.83)** | | 1.03 (0.76; 1.39) | **0.22 (0.06; 0.38)** | |
| **PA (cohort-specific quartiles)** | | | | | | | |
| 1st quartile (less activity) | 785 | Ref. | Ref. | | Ref. | Ref. | |
| 2nd quartile | 771 | Ref. | 1.16 (0.88; 1.52) | | 0.74 (0.54; 1.02) | -0.16 (-0.31; 0.00) | |
| 3rd quartile | 784 | Ref. | 0.92 (0.69; 1.21) | | 0.89 (0.64; 1.22) | 0.02 (-0.14; 0.18) | |
| 4th quartile | 701 | Ref. | 0.79 (0.59; 1.06) | * | 0.83 (0.60; 1.16) | 0.08 (-0.09; 0.25) | |
| **BMI** | | | | | | | |
| Normoweight | 705 | Ref. | Ref. | | Ref. | Ref. | |
| Overweight | 1391 | Ref. | 0.86 (0.67; 1.11) | | 0.99 (0.74; 1.32) | -0.02 (-0.17; 0.12) | |
| Obese | 945 | Ref. | 1.04 (0.78; 1.38) | | 1.05 (0.75; 1.46) | -0.12 (-0.29; 0.04) | |
| **ST (cohort-specific quartiles)**§ | | | | | | | |
| 1st quartile (less time) | 635 | Ref. | Ref. | | Ref. | Ref. | |
| 2nd quartile | 505 | Ref. | 1.03 (0.75; 1.42) | | 1.05 (0.72; 1.55) | -0.06 (-0.24; 0.12) | |
| 3rd quartile | 554 | Ref. | 1.19 (0.87; 1.63) | | 1.17 (0.81; 1.69) | -0.05 (-0.23; 0.12) | |
| 4th quartile | 518 | Ref. | 0.80 (0.57; 1.12) | | 0.95 (0.65; 1.38) | 0.05 (-0.13; 0.23) | |
| **Sleep characteristics** | | | | | | | |
| **Hours of night-time sleep** | | | | | | | |
| Normal sleep | 2208 | Ref. | Ref. | | Ref. | Ref. | |
| Short sleep (≤6 h) | 529 | Ref. | 1.05 (0.81; 1.36) | | 1.05 (0.77; 1.41) | -0.02 (-0.17; 0.13) | |
| Long sleep (≥9 h) | 304 | Ref. | **1.66 (1.22; 2.28)** | * | **0.62 (0.38; 1.00)** | **-0.51 (-0.71; -0.32)** | * |

| | n | No changes | Decreased ≥1 point | Increased ≥1 point | Mean changes |
|---|---|---|---|---|---|
| | | **Changes in MEDAS** | | | |
| | | | OR (95%CI) | OR (95%CI) | $\bar{x}$ (95%CI) |
| | | n=162 | n=949 | n=470 | n=3045 |
| **Hours of day-time sleep**‖ | | | | | |
| None | 600 | Ref. | Ref. | Ref. | Ref. |
| Short nap (≤30 min) | 691 | Ref. | 1.22 (0.90; 1.66) | 1.02 (0.72; 1.44) | -0.00 (-0.17; 0.17) |
| Long nap (30-60 min) | 350 | Ref. | 1.05 (0.76; 1.46) | 0.92 (0.60; 1.41) | 0.04 (-0.16; 0.24) |
| Very long nap (≥60min) | 100 | Ref. | 1.05 (0.51; 2.18) | 1.27 (0.66; 2.42) | 0.08 (-0.24; 0.41) |
| **Overall sleep quality**§ | | | | | |
| Very good | 430 | Ref. | 1.23 (0.91; 1.68) | **1.44 (1.00; 2.13)** | -0.02 (-0.18; 0.17) |
| Good | 1268 | Ref. | Ref. | Ref. | Ref. |
| Fair | 429 | Ref. | 1.03 (0.74; 1.43) | 1.01 (0.70; 1.45) | 0.03 (-0.14; 0.21) |
| Poor/very poor | 65 | Ref. | 1.01 (0.49; 2.10) | 0.80 (0.32; 2.02) | 0.04 (-0.36; 0.44) |
| **Health variables** | | | | | |
| **Overall health** | | | | | |
| 1st quartile (worst) | 616 | Ref. | Ref. | Ref. | Ref. |
| 2nd quartile | 556 | Ref. | 0.90 (0.65; 1.26) | 1.01 (0.69; 1.48) | 0.10 (-0.08; 0.29) |
| 3rd quartile | 622 | Ref. | 0.90 (0.65; 1.25) | 1.22 (0.84; 1.79) | 0.16 (-0.03; 0.34) |
| 4th quartile (best) | 1247 | Ref. | 0.91 (0.66; 1.25) | 1.58 (1.08; 2.31) * | 0.28 (0.09; 0.46) * |
| **Pain scale¶** | 1323 | Ref. | 1.09 (0.98; 1.20) | 1.01 (0.92; 1.11) | -0.01 (-0.06; 0.04) |
| **Chronic morbidities** | | | | | |
| Diabetes | 532 | Ref. | 0.93 (0.72; 1.21) | 1.16 (0.87; 1.55) | 0.08 (-0.07; 0.23) |
| Hypertension | 1690 | Ref. | 1.01 (0.82; 1.24) | **0.73 (0.58; 0.93)** | -0.08 (-0.20; 0.04) |
| CVD† | 199 | Ref. | 0.81 (0.54; 1.22) | 0.87 (0.55; 1.39) | 0.05 (-0.18; 0.29) |
| Cancer† | 188 | Ref. | 1.12 (0.74; 1.68) | **1.45** (0.95; 2.22) | 0.11 (-0.12; 0.35) |
| Osteomuscular disease | 1155 | Ref. | 0.89 (0.71; 1.11) | 0.93 (0.72; 1.19) | 0.05 (-0.08; 0.18) |
| Depression | 368 | Ref. | 1.12 (0.81; 1.55) | 0.92 (0.64; 1.34) | **-0.11** (-0.29; 0.08) |
| **Mobility limitations**¶ | 476 | Ref. | 1.21 (0.78; 1.88) | 0.95 (0.64; 1.43) | -0.15 (-0.37; 0.05) |
| **Negative ageing experience scale**¶ | 1313 | Ref. | 1.11 (0.93; 1.33) | 0.92 (0.78; 1.08) | -0.05 (-0.14; 0.04) |
| **Cantril ladder**‡ | 1770 | Ref. | 1.05 (0.97; 1.15) | 1.05 (0.96; 1.14) | -0.01 (-0.06; 0.04) |
| **Low MMSE scores (<23)**‡ | 77 | Ref. | 0.68 (0.33; 1.38) | 0.97 (0.43; 2.18) | -0.01 (-0.38; 0.35) |
| **Years since baseline examination** | 3041 | Ref. | 1.05 (0.86; 1.29) | 1.02 (0.81; 1.28) | -0.03 (-0.15; 0.09) |
| **Week of the state of alarm** | 3041 | Ref. | 1.09 (0.99; 1.21) | 1.00 (0.89; 1.13) | 0.00 (-0.05; 0.07) |

OR: Odds Ratio. 95%CI: 95% Confidence interval. $\bar{x}$: Mean changes. Confidence intervals are represented by the lower limit and the upper limit with either a hyphen (when the main estimates is an OR) or a semicolon (when the main estimate is a difference in means); MMSE: Mini Mental State Examination

PA: Physical activity; BMI: Body Mass Index; ST: Sedentary time

Note: Figures highlighted in bold show a statistically significant association.

†Information not available in *EXERNET*; ‡ Information only available in the *Seniors-ENRICA*-2 and *Edad con Salud* cohorts; §Information not available in the *TSHA* cohort; ‖Information only available in the *Seniors ENRICA-2* and *Exernet* cohorts; ¶ Information only available in the *Seniors-ENRICA-2* cohort

All models were adjusted for baseline age, sex (men or women), educational level (primary or less, secondary, or university), civil status (married, widowed, never married, divorced), housing conditions (lack of outdoor views, terrace/balcony, private garden/yard, lack of internet and noise), smoking status (never, former, or current), adherence to the Mediterranean diet (MEDAS score), body mass index (normoweight, overweight, or obese), physical activity (cohort-specific quartiles), hours of sleep at night (normal, short sleep, long sleep), chronic comorbidities (hypertension, diabetes, osteo-muscular disease and depression), overall health (SF-12, WHODAS-12 o EQ5D), time since last follow-up visit, cohort of study and week of the state of alarm.

* p-value for trend <0.05

**Supplementary Table 4.** Prospective association between participant characteristics at baseline and changes in weight between the COVID-19 pre-confinement period and the State of Alarm (n=1561: 792 from *Seniors-ENRICA*-2, 239 from *Edad con Salud*, 312 from *TSHA* and 218 from *Exernet*).

| | Changes in weight | | | | |
|---|---|---|---|---|---|
| | **n** | **Maintained** | **Increased ≥1 kilos** | **Decreased≥1 kilos** | **Mean changes (kg)** |
| | | | OR (95%CI) | OR (95%CI) | $\bar{x}$ (95%CI) |
| | | n=480 | n=465 | n=616 | n=1562 |
| **Socio-demographic characteristics** | | | | | |
| **Age, yr** | 1561 | 1561 | 1.00 (0.97; 1.03) | 1.01 (0.98; 1.04) | -0.01 (-0.06; 0.03) |
| **Sex** | | | | | |
| Male | 715 | Ref. | Ref. | Ref. | Ref. |
| Female | 846 | Ref. | 0.69 (0.46; 1.05) | 0.94 (0.63; 1.39) | **-0.73 (-1.35; -0.12)** |
| **Education** | | | | | |
| Primary or less | 952 | Ref. | Ref. | Ref. | Ref. |
| Secondary | 299 | Ref. | 1.04 (0.73; 1.50) | 1.06 (0.75; 1.51) | -0.03 (-0.57; 0.51) |
| University | 310 | Ref. | 0.82 (0.56; 1.19) | 1.14 (0.81; 1.63) | **-0.59 (-1.14; -0.03)** |
| **Civil status** | | | | | |
| Married | 1066 | Ref. | Ref. | Ref. | Ref. |
| Never married | 77 | Ref. | 1.04 (0.53; 2.06) | 0.79 (0.40; 1.57) | 0.03 (-1.01; 1.08) |
| Divorced | 82 | Ref. | 1.31 (0.67; 2.58) | 0.90 (0.45; 1.78) | 0.53 (-0.49; 1.55) |
| Widowed | 336 | Ref. | 0.85 (0.53; 1.37) | 0.74 (0.47; 1.17) | 0.12 (-0.60; 0.85) |
| **Living alone** | | | | | |
| No | 1210 | Ref. | Ref. | Ref. | Ref. |
| Yes | 351 | Ref. | 1.19 (0.74; 1.91) | 1.54 (0.96; 2.45) | -0.23 (-0.97; 0.48) |
| **Daily contact family/friends**† | | | | | |
| Yes | 1089 | Ref. | Ref. | Ref. | Ref. |
| No | 251 | Ref. | 0.82 (0.55; 1.22) | 1.07 (0.74; 1.56) | -0.17 (-0.77; 0.43) |
| **Feeling lonely**‡ | 1028 | Ref. | 1.02 (0.85; 1.23) | 0.93 (0.77; 1.14) | 0.13 (-0.14; 0.40) |
| **Housing conditions** | | | | | |
| **Lack outdoor views** | | | | | |
| No | 1440 | Ref. | Ref. | Ref. | Ref. |
| Yes | 121 | Ref. | 1.15 (0.70; 1.89) | 0.94 (0.58; 1.54) | 0.26 (-0.49; 1.02) |
| **Lack terrace/balcony** | | | | | |
| No | 1128 | Ref. | Ref. | Ref. | Ref. |
| Yes | 433 | Ref. | 0.85 (0.63; 1.15) | 0.82 (0.61; 1.09) | 0.24 (-0.21; 0.68) |
| **Lack garden/yard** | | | | | |
| No | 392 | Ref. | Ref. | Ref. | Ref. |
| Yes | 1169 | Ref. | 0.83 (0.59; 1.16) | 0.81 (0.59; 1.10) | 0.33 (-0.16; 0.82) |
| **Internet access** | | | | | |
| Yes | 1133 | Ref. | Ref | Ref. | Ref. |
| No | 428 | Ref. | 0.84 (0.59; 1.20) | 0.99 (0.71; 1.38) | -0.29 (-0.80; 0.23) |

| | n | Maintained | Increased ≥1 kilos | Decreased≥1 kilos | Mean changes (kg) |
|---|---|---|---|---|---|
| | | | **Changes in weight** | | |
| | | | OR (95%CI) | OR (95%CI) | $\bar{x}$ (95%CI) |
| | | n=480 | n=465 | n=616 | n=1562 |
| **Too much noise** | | | | | |
| No | 1510 | Ref. | Ref. | Ref. | Ref. |
| Yes | 51 | Ref. | 1.56 (0.71; 3.45) | 1.61 (0.76; 3.40) | 0.07 (-1.04; 1.19) |
| **Lifestyle-behaviours** | | | | | |
| **Smoking** | | | | | |
| Never smokers | 895 | Ref. | Ref. | Ref. | Ref. |
| Former smokers | 543 | Ref. | 0.99 (0.72; 1.37) | 0.97 (0.71; 1.31) | -0.23 (-0.71; 0.25) |
| Smoker | 115 | Ref. | **2.00 (1.18; 3.39)** | 0.13 (0.65; 2.00) | 0.66 (-0.15; 1.47) |
| **Alcohol intake**† | | | | | |
| Non drinker | 614 | Ref. | Ref. | Ref. | Ref. |
| Drinker, not daily | 335 | Ref. | **0.67 (0.47; 0.96)** | **0.63 (0.44; 0.90)** | 0.28 (-0.29; 0.85) |
| Drinker, daily or almost daily | 394 | Ref. | 0.77 (0.53; 1.11) | 0.73 (0.51; 1.05) | 0.22 (-0.35; 0.79) |
| **MEDAS** | 1561 | Ref. | 1.05 (0.97; 1.14) | 1.01 (0.93; 1.09) | 0.10 (-0.02; 0.21) |
| **PA (cohort-specific quartiles)** | | | | | |
| 1st quartile | 369 | Ref. | Ref. | Ref. | Ref. |
| 2nd quartile | 385 | Ref. | 1.29 (0.87; 1.92) | 0.99 (0.69; 1.52) | **0.60 (0.03; 1.17)** |
| 3rd quartile | 422 | Ref. | 1.25 (0.85; 1.86) | 0.92 (0.64; 1.32) | 0.32 (-0.25; 0.90) |
| 4th quartile | 385 | Ref. | 1.41 (0.87; 1.96) | 0.86 (0.59; 1.26) | 0.49 (-0.10; 1.08) |
| **BMI** | | | | | |
| Normoweight | 480 | Ref. | Ref. | Ref. | Ref. |
| Overweight | 465 | Ref. | 0.79 (0.52; 1.23) | 1.00 (0.66; 1.53) | 0.39 (-0.25; 1.04) |
| Obese | 616 | Ref. | 0.92 (0.42; 1.99) | 1.31 (0.64; 2.59) | 0.53 (-0.57; 1.63) |
| **ST (cohort-specific quartiles)**§ | | | | | |
| 1st quartile | 351 | Ref. | Ref. | Ref. | Ref. |
| 2nd quartile | 293 | Ref. | 1.08 (0.72; 1.63) | 1.26 (0.84; 1.89) | -0.07 (-0.63; 0.49) |
| 3rd quartile | 314 | Ref. | 1.04 (0.69; 1.56) | 1.10 (0.74; 1.65) | 0.05 (-0.50; 0.60) |
| 4th quartile | 291 | Ref. | 0.88 (0.57; 1.35) | 0.93 (0.61; 1.40) | -0.10 (-0.68; 0.48) |
| **Sleep characteristics** | | | | | |
| **Hours of night-time sleep** | | | | | |
| Normal sleep | 1168 | Ref. | Ref. | Ref. | Ref. |
| Short sleep | 259 | Ref. | 0.70 (0.48; 1.02) | 1.04 (0.75; 1.46) | **-0.67 (-1.22; -0.13)** |
| Long sleep | 134 | Ref. | 1.40 (0.83; 2.37) | 1.49 (0.91; 2.43) | -0.14 (-0.87; 0.59) |
| **Hours of day-time sleep**‖ | | | | | |
| None | 332 | Ref. | Ref. | Ref. | Ref. |
| Short nap | 433 | Ref. | 0.82 (0.55; 1.23) | 0.87 (0.58; 1.29) | -0.08 (-0.59; 0.43) |
| Long nap | 185 | Ref. | 1.06 (0.64; 1.75) | 1.06 (0.65; 1.72) | 0.11 (-0.51; 0.72) |

| | Changes in weight | | | | |
|---|---|---|---|---|---|
| | n | Maintained | Increased ≥1 kilos | Decreased≥1 kilos | Mean changes (kg) |
| | | | OR (95%CI) | OR (95%CI) | $\bar{x}$ (95%CI) |
| | | n=480 | n=465 | n=616 | n=1562 |
| Very long nap | 58 | Ref. | 0.65 (0.27; 1.54) | 1.20 (0.56; 2.59) | **-1.29 (-2.27; -0.30)** |
| **Health variables** | | | | | |
| **Overall sleep quality§** | | | | | |
| Very good | 234 | Ref. | 0.96 (0.64; 1.45) | **0.66 (0.44; 0.99)** | **0.73 (0.17; 1.29)** |
| Good | 734 | Ref. | Ref. | Ref. | Ref. |
| Fair | 235 | Ref. | 0.96 (0.63; 1.44) | 1.07 (0.72; 1.60) | 0.16 (-0.40; 0.71) |
| Poor/very poor | 33 | Ref. | 1.51 (0.51; 4.49) | 1.64 (0.61; 4.42) * | -0.10 (-1.42; 1.21) |
| **Overall health** | | | | | |
| 1st quartile | 267 | Ref. | Ref. | Ref. | Ref. |
| 2nd quartile | 306 | Ref. | 0.96 (0.61; 1.51) | 0.97 (0.62; 1.51) | 0.04 (-0.64; 0.72) |
| 3rd quartile | 339 | Ref. | 0.92 (0.58; 1.43) | 1.00 (0.65; 1.56) | -0.08 (-0.76; 0.60) |
| 4th quartile (best) | 649 | Ref. | 0.80 (0.51; 0.26) | 1.01 (0.65; 1.56) | -0.18 (-0.85; 0.50) |
| **Pain scale ¶** | 650 | Ref. | 1.04 (0.93; 1.17) | 1.00 (0.88; 1.12) | 0.09 (-0.14; 0.16) |
| **Chronic morbidities** | | | | | |
| Diabetes | 256 | Ref. | 1.07 (0.73; 1.55) | 0.93 (0.65; 1.32) | 0.03 (-0.52; 0.58) |
| Hypertension | 830 | Ref. | 0.98 (0.74; 1.30) | 0.91 (0.69; 1.19) | 0.21 (-0.21; 0.64) |
| CVD† | 92 | Ref. | 1.14 (0.62; 2.13) | 1.08 (0.60; 1.93) | **0.99 (0.07; 1.90)** |
| Cancer† | 102 | Ref. | 1.10 (0.63; 1.91) | 1.20 (0.71; 2.03) | -0.30 (-1.13; 0.52) |
| Osteomuscular | 597 | Ref. | **1.32 (0.99; 1.79)** | 1.07 (0.80; 1.54) | 0.36 (-0.09; 0.81) |
| Depression | 154 | Ref. | 1.29 (0.80; 2.11) | 1.30 (0.81; 2.08) | -0.09 (-0.79; 0.62) |
| **Mobility limitations¶** | 259 | Ref. | 1.09 (0.67; 1.79) | 1.06 (0.64; 1.77) | -0.07 (-0.71; 0.56) |
| **Negative ageing experience scale¶** | 766 | Ref. | 1.01 (0.83; 1.23) | 0.87 (0.72; 3.00) | **0.27 (0.01; 0.52)** |
| **Cantril ladder¶** | 1025 | Ref. | 0.95 (0.86; 1.05) | **0.90 (0.81; 0.99)** | 0.07 (-0.07; 0.21) |
| **Low MMSE scores (<23)¶** | 26 | Ref. | 0.91 (0.31; 2.67) | 0.80 (0.25; 2.60) | 0.70 (-0.84; 2.26) |
| **Years since baseline examination** | 1561 | Ref. | 1.06 (0.78; 1.43) | 1.21 (0.91; 1.60) | 0.01 (-0.43; 0.45) |
| **Week state alarm** | 1561 | Ref. | 0.98 (0.85; 1.13) | 0.93 (0.82; 1.08) | 0.05 (-0.16; 0.27) |

OR: Odds Ratio. 95%CI: 95% Confidence interval; MEDAS: Mediterranean Diet Assessment Score; PA: Physical activity; ST: Sedentary time; MMSE: Mini Mental State Examination

Note: Figures highlighted in bold show a statistically significant association.

†Information not available in *EXERNET*; ‡Information only available in *Seniors ENRICA-2* and *Edad con Salud*; §Information not available in *TSHA*; ‖Information only available in *Seniors ENRICA-2* and *Exernet* ¶Information only available in *Seniors-ENRICA-2*

All models were adjusted for baseline age, sex (men or women), educational level (primary or less, secondary, or university), civil status (married, widowed, single, divorced), smoking status (never, former, or current), alcohol drinking (never, former, moderate, or heavy), adherence to the Mediterranean diet (MEDAS score), weight and height, physical activity (quartiles), hours of sleep at night (normal, short sleep, long sleep), chronic comorbidities (hypertension, diabetes, cardiovascular disease, cancer, osteo-muscular disease and depression), overall health (SF-12, WHODAS-12 o EQ5D), time since last follow-up visit, cohort of study and week of the state of alarm.

* p-value for trend <0.05

**Supplementary Table 5.** Prospective association between participant characteristics and changes in overall physical activity (n=2577: 1323 from *Seniors ENRICA-2*, 829 from *TSHA*, and 425 from *EXERNET*), recreational physical and household physical activity (n=1748: 1323 from *Seniors ENRICA-2* and 425 from *EXERNET*), and the PASE score (n=829 from *TSHA*) between the COVID-19 pre-confinement period and the State of Alarm.

| | Changes in PA | | | | | Changes in PA in Seniors-ENRICA-2 and EXERNET | | Changes in PA in TSHA |
|---|---|---|---|---|---|---|---|---|
| | **n** | **Average changes** | **Unhealthier changes#** | **Healthier changes#** | **Total PA (Mets-h/wk)** | **Recreational PA (Mets-h/wk)** | **Household PA (Mets-h/wk)** | **PASE score** |
| | | | OR (95%CI) | OR (95%CI) | $\bar{x}$ (95%CI) | $\bar{x}$ (95%CI) | $\bar{x}$ (95%CI) | $\bar{x}$ (95%CI) |
| | | n=1291 | n=643 | n=643 | n=1748 | n=1748 | n=1748 | n=829 |
| **Socio-demographic characteristics** | | | | | | | | |
| **Age, yr** | 2577 | Ref. | **1.06 (1.04; 1.09)** | **0.95 (0.92; 0.97)** | **-0.61 (-0.90; -0.32)** | **-0.29 (-0.48; -0.11)** | **-0.36 (-0.57; -0.14)** | **-2.33 (-3.04; -1.62)** |
| **Sex** | | | | | | | | |
| Male | 1088 | Ref. | Ref. | Ref. | Ref. | Ref. | Ref. | Ref. |
| Female | 1489 | Ref. | **0.59 (0.44; 0.78)** | **1.34 (1.03; 1.74)** | **11.1 (7.98; 14.20)** | **-4.66 (-6.54; -2.77)** | **15.2 (12.71; 17.64)** | 6.32 (-2.41; 15.0) |
| **Education** | | | | | | | | |
| Primary or less | 1778 | Ref. | Ref. | Ref. | Ref. | Ref. | Ref, | Ref, |
| Secondary | 402 | Ref. | 1.04 (0.76; 1.44) | 0.95 (0.72; 1.28) | -1.75 (-5.05;1.56) | -0.35 (-2.42; 1.72) | -1.51 (-3.97; 0.95) | 5.25 (-5.48; 15.98) |
| University | 397 | Ref. | 1.16 (0.81; 1.62) | 1.11 (0.83; 1.49) | -0.95 (-4.40; 2.41) | -0.36 (-2.45; 2.74) | -0.73 (-3.23; 1.77) | 0.02 (-13.0; 13.1) |
| **Civil status** | | | | | | | | |
| Married | 1671 | Ref. | Ref. | Ref. | Ref. | Ref. | Ref. | Ref. |
| Never married | 144 | Ref. | 0.85 (0.50; 1.47) | 0.74 (0.44; 1.26) | -4.31 (-10.1; 1.48) | -0.89 (-4.51; 2.73) | -3.40 (-7.72; 0.92) | -1.71 (-20.2; 16.7) |
| Divorced | 106 | Ref. | 0.94 (0.50; 1.78) | 0.61 (0.34; 1.09) | -3.03 (-0.12; 3.07) | -0.93 (-4.74; 2.89) | -2.24 (-6.78; 2.31) | 1.00 (-25.1; 27.1) |
| Widowed | 656 | Ref. | 0.84 (0.57; 1.23) | 0.97 (0.69; 1.35) | 0.74 (-3.28; 4.76) | 0.27 (-2.24; 2.78) | 0.51 (-2.49; 3.51) | -2.85 (-14.4; 8.68) |
| **Living alone** | 1944 | | | | | | | |
| No | 633 | Ref. | Ref. | Ref. | Ref. | Ref. | Ref. | Ref. |
| Yes | | Ref. | 1.05 (0.71; 1.56) | 1.23 (0.87; 1.72) | -1.22 (-5.24; 2.80) | 1.20 (-1.31; 3.72) | -2.26 (-5.26; 0.75) | 2.16 (-10.1; 14.4) |
| **Daily contact with family/friends other than cohabitants†** | | | | | | | | |
| Yes | 1759 | Ref. | Ref. | Ref. | Ref. | Ref. | Ref. | Ref. |
| No | 389 | Ref. | 1.17 (0.85; 1.62) | **0.74 (0.54;0.99)** | -1.20 (-5.17; 2.77) | -1.91 (-4.58; 0.76) | 0.72 (-2.10; 3.55) | **-11.7 (-20.0; -3.53)** |
| **Feeling lonely** | 1312 | Ref. | 0.97 (0.81; 1.15) | 1.11 (0.95; 1.31) | 0.83 (-0.68; 2.34) | -2.03 (-4.70; 0.63) | **1.31 (0.23; 2.39)** | **-** |
| **Housing conditions** | | | | | | | | |
| **Lack of outdoor views** | | | | | | | | |
| No | 2391 | Ref. | Ref. | Ref. | Ref. | Ref. | Ref. | Ref. |
| Yes | 749 | Ref. | **2.34 (1.56; 3.52)** | 1.01 (0.69; 1.57) | **-7.17 (-11.4:-2.91)** | **-2.80 (-5.44: -0.12)** | **-4.43 (-7.60; -1.25)** | 6.14 (-22.7; 10.5) |
| **Lack of terrace/balcony** | | | | | | | | |
| No | 1828 | Ref. | Ref. | Ref. | . Ref | . Ref | Ref. | Ref. |
| Yes | 749 | Ref. | 1.15 (0.90; 1.47) | 0.90 (0.72; 1.13) | **-2.55 (-5.29;0.00)** | **-1.03 (-2.74; 0.68)** | -1.54 (-3.59; 0.50) | -1.97 (-9.45; 5.51) |

| | Changes in PA | | | | | Changes in PA in Seniors-ENRICA-2 and EXERNET | | Changes in PA in TSHA |
|---|---|---|---|---|---|---|---|---|
| | n | Average changes | Unhealthier changes# | Healthier changes# | Total PA (Mets-h/wk) | Recreational PA (Mets-h/wk) | Household PA (Mets-h/wk) | PASE score |
| | | | OR (95%CI) | OR (95%CI) | $\bar{x}$ (95%CI) | $\bar{x}$ (95%CI) | $\bar{x}$ (95%CI) | $\bar{x}$ (95%CI) |
| | | n=1291 | n=643 | n=643 | n=1748 | n=1748 | n=1748 | n=829 |
| **Lack of garden/yard** | | | | | | | | |
| No | 778 | Ref. | Ref. | Ref. | Ref. | Ref. | Ref. | Ref. |
| Yes | 1799 | Ref. | 0.85 (0.64; 1.12) | **0.68 (0.54; 0.87)** | -0.60 (-3.81; 2.60) | -0.66 (-2.67; 1.35) | 0.26 (-2.13; 2.66) | -6.41 (-14.0; 1.40) |
| **Internet access** | | | | | | | | |
| Yes | 1582 | Ref. | Ref. | Ref. | Ref. | Ref. | Ref. | Ref. |
| No | 995 | Ref. | 1.10 (0.85; 1.43) | 0.90 (0.71; 1.15) | -2.29 (-5.25; 0.66) | -1.52 (-3.37; 0.33) | -0.86 (-3.07; 1.34) | 0.07 (-0.72; 7.29) |
| **Too much noise** | | | | | | | | |
| No | 2506 | Ref. | Ref. | Ref. | Ref. | Ref. | Ref. | Ref. |
| Yes | 71 | Ref. | 1.11 (0.56; 2.22) | 0.99 (0.54; 1.81) | 0.13 (-7.35; 7.60) | 0.52 (-5.20; 4.17) | 0.33 (-5.25; 5.91) | -5.73 (-25.1; 13.6) |
| **Lifestyle-behaviours** | | | | | | | | |
| **Smoking** | | | | | | | | |
| Never smokers | 1593 | Ref. | Ref. | Ref. | Ref. | Ref. | Ref. | Ref. |
| Former smokers | 181 | Ref. | 1.04 (0.79; 1.39) | 0.83 (0.64; 1.07) | -1.70 (-4.60; 1.20) | 0.73 (-1.08; 2.55) | **-2.35 (-4.52; -0.19)** | -5.58 (-15.3; 4.13) |
| Smoker | 785 | Ref. | 1.06 (0.67; 1.69) | 0.71 (0.46; 1.07) | -3.44 (-8.24; 1.45) | -2.81 (-5.82; 0.19) | -0.86 (-4.44; 2.70) | -11.5 (-26.1; 3.08) |
| **Alcohol intake**† | | | | | | | | |
| Not drinker | 1079 | Ref. | Ref. | Ref. | Ref. | Ref. | Ref. | Ref. |
| Drinker, not daily | 455 | Ref. | 0.95 (0.69; 1.30) | 1.18 (0.88; 1.59) | 1.40 (-1.90; 4.70) | -0.15 (-2.36; 2.07) | 1.72 (-0.62; 4.07) | 3.75 (-8.18; 15.68) |
| Drinker, daily/almost daily | 618 | Ref. | 0.76 (0.56; 1.04) | 1.01 (0.76; 1.35) | 1.39 (-2.05; 4.82) | 1.03 (-1.27; 3.33) | 0.60 (-1.85; 3.05) | -0.36 (-9.14; 8.42) |
| **MEDAS** | 2577 | Ref. | 0.96 (0.90; 103.) | **1.05** (0.99; 1.11) | **1.33 (0.66; 2.01)** | **0.73 (0.31; 1.16)** | **0.70 (0.20; 1.20)** | 1.30 (-0.79; 3.39) |
| **BMI** | | | | | | | | |
| Normoweight | 561 | Ref. | Ref. | Ref. | Ref. | Ref. | Ref. | Ref. |
| Overweight | 1205 | Ref. | 1.06 (0.80; 1.43) | 0.94 (0.72; 1.22) | 0.37 (-2.56; 3.30) | 1.03 (-0.81; 2.86) | -0.54 (-2.73; 1.64) | -7.45 (-18.0; 3.13) |
| Obese | 810 | Ref. | **1.37 (1.00; 1.91)** | 0.96 (0.72; 1.30) | -2.89 (-6.39; 0.62) | -1.47 (-3.66; 0.72) | -1.47 (-4.08; 1.15) | -6.30 (-17.1; 4.48) |
| **ST (cohort-specific quartiles)**‡ | | | | | | | | |
| 1st quartile (less time) | 477 | Ref. | Ref. | Ref. | Ref. | Ref. | Ref. | - |
| 2nd quartile | 417 | Ref. | 1.22 (0.84; 1.78) | 1.06 (0.76; 1.49) | 0.22 (-3.09; 3.54) | 0.40 (-1.67; 2.47) | -0.13 (-2.61; 2.34) | - |
| 3rd quartile | 440 | Ref. | 0.97 (0.67; 1.41) | 0.84 (0.60; 1.17) | -0.74 (-4.02; 2.54) | -0.44 (-1.61; 2.49) | -1.24 (-3.69; 1.21) | - |
| 4th quartile | 414 | Ref. | 0.94 (0.64; 1.39) | **0.67 (0.47; 0.96)** | -2.60 (-5.98; 0.77) | -1.00 (-3.11; 1.11) | -1.62 (-4.14; 0.90) | - |
| **Sleep characteristics** | | | | | | | | |
| **Hours night-time sleep** | | | | | | | | |
| Normal sleep | 1850 | Ref. | Ref. | Ref. | Ref. | Ref. | Ref. | Ref. |
| Short sleep (≤6 h) | 474 | Ref. | 1.07 (0.80; 1.44) | 0.87 (0.67; 1.13) | -2.82 (-5.91; 0.27) | -0.48 (-2.42; 1.45) | **-2.39 (-4.69; -0.09)** | **11.5 (2.25; 20.7)** |
| Long sleep (≥9 h) | 253 | Ref. | 1.09 (0.74; 1.60) | **0.57 (0.39; 0.84)** | -4.68 (-9.99; 0.62) | -2.50 (-5.82; 0.82) | -2.25 (-6.21; 1.71) | -5.89 (-14.9; 3.20) |

| | n | Changes in PA | | | | Changes in PA in Seniors-ENRICA-2 and EXERNET | | Changes in PA in TSHA |
|---|---|---|---|---|---|---|---|---|
| | | Average changes | Unhealthier changes# | Healthier changes# | Total PA (Mets-h/wk) | Recreational PA (Mets-h/wk) | Household PA (Mets-h/wk) | PASE score |
| | | | OR (95%CI) | OR (95%CI) | $\bar{x}$ (95%CI) | $\bar{x}$ (95%CI) | $\bar{x}$ (95%CI) | $\bar{x}$ (95%CI) |
| | | n=1291 | n=643 | n=643 | n=1748 | n=1748 | n=1748 | n=829 |
| **Hours day-time sleep**§ | | | | | | | | |
| None | 600 | Ref. | Ref. | Ref. | Ref. | Ref. | Ref. | . |
| Short nap (≤30 min) | 691 | Ref. | 0.98 (0.71; 1.36) | 1.25 (0.93; 1.68) | 0.88 (-1.98; 3.74) | 2.32 (0.53; 4.11) | -1.38 (-3.51; 0.76) | - |
| Long nap (30-60 min) | 350 | Ref. | 1.25 (0.85; 1.83) | 1.01 (0.71; 1.44) | -2.57 (-5.93; 0.79) | -0.38 (-2.48; 1.72) | -2.11 (-4.62; 0.39) | - |
| Very long (≥60min) | 100 | Ref. | 0.96 (0.49; 1.91) | 1.04 (0.59; 1.81) | -2.77 (-8.23; 2.69) | -0.37 (-3.78; 3.04) | -2.54 (-6.61; 1.53) | - |
| **Overall sleep quality**‡ | | | | | | | | |
| Very good | 236 | Ref. | 1.29 (0.85; 1.75) | 0.86 (0.59; 1.27) | -3.19 (-6.82; 0.45) | **-2.54 (-4.81; -0.25**) | -0.51 (-3.23; 2.21) | - |
| Good | 1048 | Ref. | Ref. | Ref. | Ref. | Ref. | Ref. | - |
| Fair | 383 | Ref. | 0.93 (0.65; 1.33) | 1.16 (0.84; 1.59) | 2.96 (-0.14; 6.07) | 0.82 (-1.13; 2.77) | 2.10 (-0.22; 4.42) | - |
| Poor/very poor | 61 | Ref. | 0.59 (0.26; 1.36) | 0.55 (0.25; 1.20) | 2.75 (-4.23; 9.73) * | 1.45 (-2.92; 5.84) * | 1.02 (-4.19; 6.24) | - |
| **Health variables** | | | | | | | | |
| **Overall health** | | | | | | | | |
| 1st quartile (worst) | 499 | Ref. | Ref. | Ref. | Ref. | Ref. | Ref. | Ref. |
| 2nd quartile | 475 | Ref. | 0.88 (0.60; 1.30) | 1.02 (0.73; 1.43) | **6.08 (2.47; 9.69)** | 1.47 (-0.79; 3.73) | **4.68 (1.98; 7.37)** | -8.06 (-25.4; 9.25) |
| 3rd quartile | 568 | Ref. | 1.08 (0.74; 1.57) | 1.22 (0.87; 1.70) | **5.15 (1.43; 8.87)** | 2.51 (0.18; 4.84) | **2.99 (0.23; 5.76)** | 7.75 (-7.43; 22.92) |
| 4th quartile (best) | 1035 | Ref. | 0.81 (0.54; 1.20) | 1.20 (0.85; 1.66) | **6.67 (2.77; 10.58)** | 3.09 (0.64; 5.54) | **4.06 (1.16; 6.96)** | 6.98 (-7.17; 21.1) |
| **Pain scale** ¶ | 1049 | Ref. | 1.00 (0.91; 1.09) | 0.94 (0.86; 1.03) | -0.10 (-0.89; 0.69) | -0.33 (-0.86; 0.21) | 0.20 (-0.36; 0.76) | - |
| **Chronic morbidities** | | | | | | | | |
| Diabetes | 458 | Ref. | 1.04 (0.78; 1.40) | **0.75 (0.57; 0.99)** | **-4.48 (-7.69; -1.27)** | **-4.33 (-6.34; -2.32)** | -0.19 (-2.58; 2.21) | -3.36 (-12.1; 5.30) |
| Hypertension | 1504 | Ref. | 1.10 (0.87; 1.40) | 1.15 (0.93; 1.43) | 0.59 (-1.90; 3.08) | 0.55 (-1.00; 2.11) | -0.06 (-1.92; 1.79) | 0.30 (-7.49; 8.09) |
| CVD† | 144 | Ref. | 1.05 (0.63; 1.74) | 1.13 (0.73; 1.74) | 0.23 (-6.39; 6.84) | -2.22 (-6.66; 2.21) | 2.13 (-2.58; 6.84) | 4.65 (-6.64; 15.94) |
| Cancer† | 144 | Ref. | 1.23 (0.77; 1.97) | 0.99 (0.63; 1.53) | -5.04 (-10.8; 0.72) | -3.68 (-7.54; 0.19) | -1.12 (-5.23; 2.98) | 2.30 (-9.89; 10.62) |
| Osteomuscular disease | 965 | Ref. | 0.94 (0.73; 1.21) | 1.15 (0.92; 1.44) | **3.64 (0.02; 6.27)** | **2.15 (0.51; 3.79)** | 1.66 (-0.29; 3.62) | -2.70 (-11.0; 5.65) |
| Depression | 324 | Ref. | 1.22 (0.83; 1.79) | 0.90 (0.65; 1.24) | -1.12 (-5.02; 2.79) | -1.89 (-4.44; 0.55) | 0.49 (-2.42; 3.39) | **-12.6 (-24.0; -1.12)** |
| **Mobility limitations**¶ | 476 | Ref. | 1.27 (0.86; 1.89) | 1.10 (0.76; 1.60) | -1.22 (-4.66; 2.22) | 0.00 (-2.30; 2.30) | -1.93 (-4.36; 0.51) | - |
| **Negative ageing experience scale**¶ | 1313 | Ref. | 0.98 (0.83; 1.14) | 0.99 (0.84; 1.16) | 0.52 (-0.92; 1.95) | -0.16 (-0.12; 0.80) | 0.62 (-0.39; 1.64) | - |
| **Cantril ladder**¶ | 1313 | Ref. | 1.09 (0.99; 1.20) | 1.01 (0.93; 1.10) | -0.45 (-1.23;0.32) | 0.12 (-0.40; 0.64) | -0.62 (-1.17; -0.07) | - |
| **Low MMSE scores (<23)** ¶ | 45 | Ref. | 1.10 (0.45-2.71) | 0.56 (0.22-1.42) | -5.72 (-13.5;2.06) | -2.13 (-7.36; 3.10) | -3.47 (-9.03; 2.08) | **-** |
| **Years since baseline exam** | 2577 | Ref. | 1.17 (0.94; 1.12) | 1.10 (0.90; 1.33) | 0.89 (-1.77; 3.55) | -1.89 (-4.34; 0.55) | -0.15(-2.13; 1.84) | -2.31 (-7.57; 2.95) |
| **Week of the state of alarm** | 2577 | Ref. | 0.99 (0.88; 1.45) | **1.17 (1.04; 1.30)** | **1.45 (0.24; 2.66)** | **1.27 (0.51; 2.02)** | **0.19 (-0.71; 1.10)** | **8.83 (4.31; 13.4)** |

OR: Odds Ratio. 95%CI: 95% Confidence interval. $\bar{x}$: Mean change. Confidence intervals are represented by the lower limit and the upper limit with either a hyphen (when the main estimates is an OR) or a semicolon (when the main estimate is a difference in means)

**#**Unhealthier changes were defined by decreasing physical activity more than the observed 75th percentile change. Healthier changes were defined by decreasing physical activity less than observed 25th percentile change or even increasing physical activity with confinement
MEDAS: Mediterranean Diet Assessment Score; BMI: Body Mass Index; ST: Sedentary time; MMSE: Mini Mental State Examination.
Note: Figures highlighted in bold show a statistically significant association.
†Information not available in *EXERNET*; ‡ Information not available in *TSHA*; §Information only available in *Seniors ENRICA-2* and *Exernet*; ¶Information only available in *Seniors-ENRICA-2*
All models were adjusted for baseline age, sex (men or women), educational level (primary or less, secondary, or university), civil status (married, widowed, never married, divorced), smoking status (never, former, or current), alcohol drinking (never, former, moderate, or heavy), adherence to the Mediterranean diet (MEDAS score), body mass index (normoweight, overweight, or obese), physical activity (quartiles), hours of sleep at night (normal, short sleep, long sleep), chronic comorbidities (hypertension, diabetes, cardiovascular disease, cancer, osteo-muscular disease and depression), overall health (SF-12, WHODAS-12 or EQ5D), time since last follow-up visit, cohort of study and week of the state of alarm.
* p-value for trend <0.05

**Supplementary table 6.** Prospective association between participant characteristics at baseline and changes in overall sedentary time (n=1746: 1321 from Seniors-ENRICA-2 and 425 from Exernet), as well as in specific sedentary activities (n=1270 for TV viewing, 1196 for other screen time and 1297 for reading time in participants from the Seniors-ENRICA-2) between the COVID-19 pre-confinement period and the State of Alarm.

| | | Changes in sedentary time | | | | Changes in sedentary time (hours /day) in Seniors-ENRICA-2 | | |
|---|---|---|---|---|---|---|---|---|
| | n | Average changes | Unhealthier changes# | Healthier changes# | Mean changes overall sedentary time (hours/day) | Mean changes TV viewing time | Mean changes other screen time | Mean changes reading time |
| | | | OR (95%CI) | OR (95%CI) | $\bar{x}$ (95%CI) | $\bar{x}$ (95%CI) | $\bar{x}$ (95%CI) | $\bar{x}$ (95%CI) |
| | | n=880 | n=431 | n=437 | n=1748 | n=1270 | n=1196 | n=1297 |
| **Socio-demographic characteristics** | | | | | | | | |
| **Age, yr** | 1746 | Ref. | **0.96 (0.93;0.99)** | **1.04 (1.01;1.08)** | **-0.07 (-0.11; -0.04)** | **-0.03 (-0.06; -0.00)** | **-0.05 (-0.07; -0.02)** | 0.01 (-0.00; 0.02) |
| **Sex** | | | | | | | | |
| Male | 757 | Ref. | Ref. | Ref. | Ref. | Ref. | Ref. | Ref. |
| Female | 989 | Ref. | 1.09 (0.79;1.51) | **1.54 (1.08;2.19)** | **-0.47 (-0.85; -0.10)** | **-0.31 (-0.60; -0.02)** | 0.08 (-0.14; 0.30) | **0.19 (0.03; 0.45)** |
| **Education** | | | | | | | | |
| Primary or less | 1118 | Ref. | Ref. | Ref. | Ref. | **Ref.** | Ref. | Ref. |
| Secondary | 299 | Ref. | 1.14 (0.81;1.61) | 0.98 (0.68;1.40) | 0.03 (-0.36; 0.43) | -0.28 (-0.58; 0.02) | 0.14 (-0.08; 0.36) | -0.06 (-0.58; 0.02) |
| University | 329 | Ref. | 1.13 (0.80;1.60) | 0.72 (0.49;1.05) | **0.44 (0.04; 0.85)** | **-0.50 (-0.78; -0.19)** | **0.36 (0.15; 0.57)** | 0.08 (-0.08; 0.26) |
| **Civil status** | | | | | | | | |
| Married | 1107 | Ref. | Ref. | Ref. | Ref. | Ref, | Ref. | Ref. |
| Never married | 109 | Ref. | 0.81 (0.43;1.52) | 0.95 (0.50;1.79) | -0.06 (-0.75; 0.64) | 0.15 (-0.37; 0.67) | 0.23 (-0.18; 0.63) | -0.14 (-0.43; 0.15) |
| Divorced | 89 | Ref. | 1.32 (0.73;2.40) | 0.67 (0.33;1.38) | 0.63 (-0.10; 1.37) | **0.84 (0.30; 1.37)** | 0.07 (-0.33; 0.46) | -0.07 (-0.37; 0.23) |
| Widowed | 441 | Ref. | 0.90 (0.58;1.39) | 0.81 (0.52;1.25) | 0.08 (-0.40; 0.57) | 0.19 (-0.20; 0.57) | 0.13 (-0.16; 0.42) | -0.02 (-0.23; 0.19) |
| **Living alone** | | | | | | | | |
| No | 1289 | Ref. | Ref. | Ref. | Ref. | Ref. | Ref. | Ref. |
| Yes | 457 | Ref. | 1.09 (0.71;1.67) | 0.88 (0.57;1.37) | 0.26 (-0.23; 0.74) | -0.21 (-0.60; 0.18) | 0.03 (-0.27; 0.33) | 0.06 (-0.15; 0.27) |
| **Daily contact** † | | | | | | | | |
| Yes | 1147 | Ref. | Ref. | Ref. | Ref. | Ref. | Ref. | Ref. |
| No | 171 | Ref. | 1.04 (0.66;1.64) | 0.89 (0.58;1.37) | 0.10 (-0.38; 0.58) | -0.14 (-0.47; 0.19) | 0.05 (-0.20; 0.29) | -0.02 (-0.21; 0.16) |
| **Feeling lonely** ‡ | **1312** | Ref. | 0.89 (0.75;1.06) | 0.92 (0.78;1.09) | -0.05 (-0.23; 0.13) | -0.02 (-0.15; 0.10) | 0.00 (-0.09; 0.10) | -0.03 (-0.10; 0.04) |
| **Housing conditions** | | | | | | | | |
| **Lack of outdoor views** | | | | | | | | |
| No | 1596 | Ref. | Ref. | Ref. | Ref. | Ref. | Ref. | Ref. |
| Yes | 150 | Ref. | 1.07 (0.70;1.63) | 0.88 (0.44;1.44) | 0.05 (-0.46; 0.56) | **0.88 (0.15; 1.59)** | 0.18 (-0.12; 0.48) | -0.05 (-0.28; 0.17) |
| **Lack terrace/balcony** | | | | | | | | |
| No | 1305 | Ref. | Ref. | Ref. | Ref. | Ref. | Ref. | Ref. |
| Yes | 441 | Ref. | 0.85 (0.64;1.14) | 1.09 (0.82;1.47) | -0.20 (-0.53; 1.13) | -0.05 (-0.30; 0.20) | 0.01 (-0.10; 0.29) | -0.09 (-0.23; 0.05) |

| | n | Changes in sedentary time | | | Changes in sedentary time (hours /day) in Seniors-ENRICA-2 | | | |
|---|---|---|---|---|---|---|---|---|
| | | **Average changes** | **Unhealthier changes#** | **Healthier changes#** | **Mean changes overall sedentary time (hours/day)** | **Mean changes TV viewing time** | **Mean changes other screen time** | **Mean changes reading time** |
| | | | OR (95%CI) | OR (95%CI) | x̄ (95%CI) | x̄ (95%CI) | x̄ (95%CI) | x̄ (95%CI) |
| | | n=880 | n=431 | n=437 | n=1748 | n=1270 | n=1196 | n=1297 |
| **Lack garden/yard** | | | | | | | | |
| No | 295 | Ref. | Ref. | Ref. | Ref. | Ref. | Ref. | Ref. |
| Yes | 1451 | Ref. | 1.29 (0.92;1.80) | 0.81(0.58;1.17) | **0.37** (-0.01; 0.76) | **0.35 (0.03; 0.68)** | **0.18 (-0.00; 0.43)** | 0.08 (-0.10; 0.26) |
| **Internet access** | | | | | | | | |
| Yes | 1206 | Ref. | Ref. | Ref. | Ref. | Ref. | Ref. | Ref. |
| No | 540 | Ref. | 0.89 (0.66;1.22) | 0.71 (0.29;1.81) | **-0.50 (-0.85; -0.13)** | 0.28 (-0.01; 0.58) | **-0.57 (-0.82; -0.32)** | -0.01 (0.45; 0.36) |
| **Too much noise** | | | | | | | | |
| No | 1701 | Ref. | Ref. | Ref. | Ref. | Ref. | Ref. | Ref. |
| Yes | 45 | Ref. | 1.61 (0.81 3.22) | **1.50 (1.08;2.07)** | 0.77 (-0.13; 1.67) | 0.02 (-0.38; 0.42) | 0.23 (-0.29; 0.74) | -0.04 (0.45; 0.37) |
| **Lifestyle-behaviours** | | | | | | | | |
| **Smoking** | | | | | | | | |
| Never smokers | 1013 | Ref. | Ref. | Ref. | Ref. | Ref. | Ref. | Ref. |
| Former smokers | 591 | Ref. | 1.04 (0.77;1.40) | 0.99 (0.72;1.37) | 0.10 (-0.25; 0.45) | 0.09 (-0.32; 0.49) | 0.05 (-0.14; 0.24) | 0.07 (-0.07; 0.21) |
| Smoker | 125 | Ref. | 1.31 (0.80;2.15) | 0.73 (0.43;1.26) | 0.43 (-0.15; 1.01) | -0.04 (-0.29; 0.22) | 0.09 (-0.02; 0.28) | 0.11 (-0.11; 0.33) |
| **Alcohol intake†** | | | | | | | | |
| Non drinker | 562 | Ref. | Ref. | Ref. | Ref. | Ref. | Ref. | Ref. |
| Drinker, not daily | 370 | Ref. | 0.94 (0.65;1.36) | 1.06 (0.74;1.52) | -0.08 (-0.48; 0.32) | -0.07 (-0.35; 0.20) | 0.04 (-0.17; 0.24) | 0.12 (-0.04; 0.26) |
| Drinker, daily/almost daily | 389 | Ref. | 0.93 (0.64;1.37) | 0.83 (0.57;1.22) | 0.06 (-0.35; 0.48) | 0.00 (-0.29; 0.29) | -0.06 (-0.27; 0.15) | 0.03 (-0.13; 0.18) |
| **MEDAS** | 1746 | Ref. | **0.93 (0.87;0.99)** | 0.94 (0.87;1.01) | -0.03 (-0.11; 0.06) | -0.03 (-0.10; 0.03) | 0.00 (-0.04; 0.05) | 0.00 (-0.03; 0.05) |
| **PA (cohort-specific quartiles)** | | | | | | | | |
| 1st quartile | 462 | Ref. | Ref. | Ref. | Ref. | **Ref.** | Ref. | Ref. |
| 2nd quartile | 444 | Ref. | 1.00 (0.72;1.41) | 1.22 (0.84;1.78) | -0.30 (-0.70; 0.10) | -0.29 (-0.60; 0.02) | -0.03 (-0.26; 0.20) | 0.12 (-0.05; 0.30) |
| 3rd quartile | 457 | Ref. | 0.80 (0.57;1.14) | 1.30 (0.89;1.91) | -0.42 (-0.83; 0.01) | **-0.33 (-0.65;-0.00)** | -0.01 (-0.25; 0.23) | -0.01 (-0.19; 0.17) |
| 4th quartile | 383 | Ref. | 0.71 (0.48;1.04) * | 1.17 (0.78;1.77) | -0.55 (-0.97; 0.09) * | -0.32 (-0.66; 0.03) | -0.20 (-0.46; 0.06) | -0.04 (-0.23; 0.15) |
| **BMI** | | | | | | | | |
| Normoweight | 444 | Ref. | Ref. | Ref. | Ref. | Ref. | Ref. | Ref. |
| Overweight | 850 | Ref. | 1.33 (0.98;1.81) | 0.93 (0.68;1.29) | 0.19 (-0.16; 0.54) | 0.08 (-0.18; 0.35) | 0.13 (-0.07; 0.32) | -0.08 (-0.23; 0.06) |
| Obese | 452 | Ref. | 1.32 (0.91;1.92) | 0.99 (0.67;1.45) | 0.15 (-0.27; 0.58) | 0.30 (-0.03; 0.63) | **0.26 (0.02; 0.51)** | -0.16 (-0.34; 0.00) |
| **Sleep characteristics** | | | | | | | | |
| **Hours night-time sleep‡** | | | | | | | | |
| Normal sleep | 1329 | Ref. | Ref. | Ref. | Ref. | Ref. | Ref. | Ref. |
| Short sleep (≤6 h) | 324 | Ref. | 1.03 (0.75;1.42) | 0.90 (0.64;1.26) | 0.16 (-0.21; 0.54) | -0.06 (-0.36; 0.24) | -0.04 (-0.26; 0.18) | **-0.17 (-0.37; 0.00)** |
| Long sleep (≥9 h) | 93 | Ref. | 1.02 (0.60;1.74) | 0.71 (0.38;1.34) | 0.33 (-0.31; 0.96) | -0.02 (-0.55; 0.52) | -0.08 (-0.50; 0.34) | 0.22 (-0.07; 0.51) |

| | n | Changes in sedentary time | | | Changes in sedentary time (hours /day) in Seniors-ENRICA-2 | | | |
|---|---|---|---|---|---|---|---|---|
| | | **Average changes** | **Unhealthier changes#** | **Healthier changes#** | **Mean changes overall sedentary time (hours/day)** | **Mean changes TV viewing time** | **Mean changes other screen time** | **Mean changes reading time** |
| | | | OR (95%CI) | OR (95%CI) | $\bar{x}$ (95%CI) | $\bar{x}$ (95%CI) | $\bar{x}$ (95%CI) | $\bar{x}$ (95%CI) |
| | | n=880 | n=431 | n=437 | n=1748 | n=1270 | n=1196 | n=1297 |
| **Hours day-time sleep** | | | | | | | | |
| None | 599 | Ref. | Ref. | Ref. | Ref. | Ref. | Ref. | Ref. |
| Short nap (≤30 min) | 691 | Ref. | 1.53 (1.13;2.07) | 0.85 (0.62;1.16) | **0.55 (0.22; 0.90)** | 0.03 (-0.23; 0.29) | 0.07 (-0.09; 0.23) | 0.07 (-0.09; 0.23) |
| Long nap (30-60 min) | 349 | Ref. | 1.15 (0.81;1.66) | 0.72 (0.50;1.05) | **0.58 (0.17; 0.98)** | 0.17 (-0.16; 0.50) | 0.11 (-0.10; 0.32) | 0.11 (-0.10; 0.32) |
| Very long nap (≥60min) | 100 | Ref. | 1.58 (0.89;2.82) | **0.52 (0.28;0.96)** * | **1.06 (0.41; 1.72)** * | 0.40 (-0.12; 0.92) | 0.08 (-0.25; 0.40) | 0.08 (-0.25; 0.40) |
| **Overall sleep quality** | | | | | | | | |
| Very good | 235 | Ref. | 1.13 (0.79;1.63) | 0.98 (0.65;1.48) | -0.04 (-0.47; 0.40) | 0.07 (-10.9; 0.33) | 0.05 (-0.32; 0.41) | -0.08 (-0.31; 0.14) |
| Good | 1047 | Ref. | Ref. | Ref. | Ref. | Ref. | Ref. | Ref. |
| Fair | 383 | Ref. | 0.70 (0.50;0.99) | 0.95 (0.68;1.33) | -0.30 (-0.67; 0.07) | 0.04 (-0.16; 0.25) | 0.13 (-0.16; 0.42) | -0.21(-0.38; -0.03) |
| Poor/very poor | 61 | Ref. | 1.12 (0.56;2.23) | 0.51 (0.22;1.19) | 0.26 (-0.58; 1.10) | 0.04 (-0.52; 0.59) | -0.11 (-0.90; 0.68) | -0.21 (-0.70; 0.27) |
| **Health variables** | | | | | | | | |
| **Overall health** | | | | | | | | |
| 1st quartile (worst) | 422 | Ref. | Ref. | Ref. | Ref. | Ref. | Ref. | Ref. |
| 2nd quartile | 415 | Ref. | 1.39 (0.94;2.06) | 1.05 (0.72;1.54) | 0.13 (-0.30; 0.57) | -0.09 (-0.43; 0.25) | -0.11 (-0.36; 0.14) | 0.19 (0.01; 0.38) |
| 3rd quartile | 444 | Ref. | 1.23 (0.83;1.84) | 0.88 (0.59;1.31) | 0.28 (-0.17; 0.72) | -0.12 (-0.47; 0.23) | 0.00 (-0.25; 0.27) | **0.17 (-0.00; 0.36)** |
| 4th quartile (best) | 465 | Ref. | 1.17 (0.77;1.77) | 0.74 (0.49;1.13) | 0.26 (-0.21; 0.73) | -0.20 (-0.58; 0.18) | 0.14 (-0.14; 0.41) | **0.27 (-0.07; 0.48)** |
| **Pain scale‡** | 1047 | Ref. | 1.04 (0.95;1.14) | 0.96 (0.88;1.04) | 0.03 (-0.07; 0.13) | **0.09 (0.02; 0.15)** | -0.01 (-0.06; 0.04) | -0.02 (-0.05; 0.02) |
| **Chronic morbidities** | | | | | | | | |
| Diabetes | 300 | Ref. | 1.06 (0.76;1.49) | **0.70 (0.49;0.99)** | 0.30 (-0.08; 0.69) | **0.40 (0.11; 0.70)** | **0.23 (0.00; 0.46)** | **-0.21 (-0.37; -0.04)** |
| Hypertension | 922 | Ref. | 1.09 (0.84;1.41) | 1.01 (0.77;1.33) | 0.09 (-0.20; 0.39) | -0.04 (-0.27; 0.19) | -0.15 (-0.32; 0.02) | 0.09 (-0.03; 0.22) |
| CVD† | 56 | Ref. | 1.67 (0.81;3.48) | 1.07 (0.53;2.16) | 0.55 (-0.25; 1.35) | 0.02 (-0.53; 0.57) | 0.01 (-0.41; 0.44) | -0.00 (-0.31; 0.30) |
| Cancer† | 72 | Ref. | 0.86 (0.42;1.74) | 1.50 (0.82;2.74) | -0.62 (-1.32; 0.08) | 0.06 (-0.44; 0.55) | **-0.43 (-0.78; -0.08)** | 0.01 (-0.25; 0.38) |
| Osteomuscular disease | 753 | Ref. | **0.75 (0.57;0.99)** | 1.12 (0.84;1.49) | -0.30 (-0.62; 0.01) | 0.01 (-0.18; 0.35) | -0.06 (-0.24; 0.12) | -0.17 (-0.38; 0.04) |
| Depression | 220 | Ref. | 0.96 (0.62;1.48) | 1.41 (0.94;2.12) | -0.38 (-0.85; 0.09) | -0.23 (-0.61; 0.16) | -0.00 (-0.30; 0.29) | -0.04 (-0.18; 0.11) |
| **Mobility limitations‡** | 475 | Ref. | 0.72 (0.48;1.06) | 0.75 (0.51;1.09) | 0.09 (-0.33; 0.51) | **0.38 (0.10; 0.67)** | 0.08 (-0.14; 0.29) | -0.00 (-0.16; 0.16) |
| **Negative ageing experience scale‡** | 1311 | Ref. | 0.91 (0.77;1.07) | 0.97 (0.83;1.14) | -0.08 (-0.25; 0.10) | 0.00 (-0.11; 0.13) | -0.05 (-0.14; 0.04) | -0.02 (-0.09; 0.04) |
| **Cantril ladder‡** | 1311 | Ref. | 0.98 (0.90;1.07) | 1.04 (0.95;1.13) | -0.05 (-0.14; 0.05) | -0.04 (-01.0; 0.03) | 0.02 (-0.03; 0.07) | -0.01 (-0.05; 0.03) |
| **Low MMSE (<23) ‡** | 45 | Ref. | 0.42 (0.13;1.33) | 0.79 (0.35;1.81) | -0.50 (-1.45; 0.44) | -0.10 (-0.76; 0.54) | -0.41 (-1.03; 0.20) | -0.16 (-0.53; 0.20) |
| **Years since exam** | 1746 | Ref. | 0.96 (0.73;1.26) | 0.78 (0.58;1.05) | -0.07 (-0.39; 0.25) | -0.01 (-0.28; 0.25) | -0.02 (0.21; 0.18) | -0.02 (-0.08; 0.04) |
| **Week of state alarm** | 1746 | Ref. | **0.89 (0.78;1.00)** | 1.12 (0.99;1.27) | **-0.30 (-0.44; -0.15)** | **-0.12 (-0.23;-0.00)** | **-0.10 (-0.19; -0.02)** | **-0.21(-0.37; -0.04)** |

OR: Odds Ratio. 95%CI: 95% Confidence interval. $\bar{x}$: Mean change. Confidence intervals are represented by the lower limit and the upper limit with either a hyphen (when the main estimates is an OR) or a semicolon (when the main estimate is a difference in means)

MEDAS: Mediterranean Diet Assessment Score; BMI: Body Mass Index; PA: Physical activity; MMSE: Mini Mental State Examination.

Note: Figures highlighted in bold show a statistically significant association.
**#**Unhealthier changes were defined by increasing sedentary time more than the observed 75th percentile change. Healthier changes were defined by increasing sedentary time less than observed 25th percentile change
‡Information not available in *EXERNET*; ‡Information only available in *Seniors ENRICA-2*; §. Information only available in *Seniors ENRICA*-2 and *Edad con Salud* cohorts.
All models were adjusted for baseline age, sex (men or women), educational level (primary or less, secondary, or university), civil status (married, widowed, never married, divorced), smoking status (never, former, or current), alcohol drinking (never, former, moderate, or heavy), adherence to the Mediterranean diet (MEDAS score), body mass index (normoweight, overweight, or obese), physical activity (quartiles), hours of sleep at night (normal, short sleep, long sleep), chronic comorbidities (hypertension, diabetes, cardiovascular disease, cancer, osteo-muscular disease and depression), overall health (SF-12, WHODAS-12 o EQ5D),time since last follow-up visit, cohort of study and week of the state of alarm.
*p-value for trend <0.05

**Supplementary table 7** Prospective association between participant characteristics and changes in night-time sleep (n=2867, 1294 from *Seniors-ENRICA-2*, 411 from *Edad con Salud*, 763 from *TSHA* and 399 from *Exernet*), sleep quality (n=2095; 1289 from *Seniors-ENRICA-2*, 430 from *Edad con Salud*, and 376 from *Exernet*), and nº of poor sleep quality indicators (n=1285 in *Seniors-ENRICA 2*) during the COVID-19 confinement.

| | Changes in night-time sleep | | | | Changes in sleep quality | | | | | |
|---|---|---|---|---|---|---|---|---|---|---|
| | n | No changes | Worsening # | Improvement # | n | No changes | Worsening ## | Improvements ## | n | Changes in nº of poor sleep indicators### |
| | | | OR (95%CI) | OR (95%CI) | | | OR (95%CI) | OR (95%CI) | | $\bar{x}$ (95%CI) |
| | | n=971 | n=2020 | n=802 | | n=1180 | n=282 | n=633 | | n=1285 |
| **Socio-demographic characteristics** | | | | | | | | | | |
| **Age, yr** | 2867 | Ref. | 1.00 (0.98; 1.02) | 1.00 (0.98; 1.02) | 2095 | Ref. | 0.99 (0.96; 1.02) | 1.01 (0.98; 1.03) | 1285 | 0.00 (-0.01; 0.02) |
| **Sex** | | | | | | | | | | |
| Male | 1225 | Ref. | Ref. | Ref. | 906 | Ref. | Ref. | Ref | 645 | Ref. |
| Female | 1642 | Ref. | 0.93 (0.74; 1.18) | 1.06 (0.82; 1.37) | 1189 | Ref. | 1.26 (0.88; 1.79) | 0.85 (0.66; 1.10) | 640 | 0.11 (-0.05; 0.27) |
| **Education** | | | | | | | | | | |
| Primary or less | 1894 | Ref. | Ref. | Ref. | 1289 | Ref. | Ref. | Ref. | 746 | Ref. |
| Secondary | 511 | Ref. | 0.89 (0.68; 1.17) | 1.15 (0.87; 1.53) | 413 | Ref. | 0.79 (0.54; 1.15) | 0.96 (0.73; 1.25) | 249 | 0.00 (-0.16; 0.17) |
| University | 462 | Ref. | 0.81 (0.60; 1.09) | 1.10 (0.80; 1.50) | 393 | Ref. | 0.78 (0.51; 1.18) | 0.93 (0.70; 1.24) | 290 | -0.06 (-0.23; 0.11) |
| **Civil status** | | | | | | | | | | |
| Married | 1876 | Ref. | Ref; | Ref. | 1340 | Ref. | Ref. | Ref. | 839 | Ref. |
| Never married | 155 | Ref. | 0.94 (0.57; 1.56) | 1.02 (0.61; 1.71) | 124 | Ref. | 0.49 (0.22; 1.09) | **1.64 (1.00; 2.67)** | 88 | 0.05 (-0.26; 0.35) |
| Divorced | 149 | Ref. | 1.01 (0.61; 1.67) | 1.20 (0.72; 1.99) | 134 | Ref. | 0.99 (0.53; 1.85) | 1.43 (0.89; 2.29) | 77 | 0.02 (-0.29; 0.33) |
| Widowed | 687 | Ref. | 1.13 (0.81; 1.56) | 1.02 (0.72; 1.44) | 497 | Ref. | 0.83 (0.52; 1.34) | 1.32 (0.93; 1.88) | 281 | 0.11 (-0.11; 0.33) |
| **Living alone** | | | | | | | | | | |
| No | 2188 | Ref. | Ref. | Ref. | 1562 | Ref. | Ref. | Ref. | 977 | Ref. |
| Yes | 679 | Ref. | 0.94 (0.67; 1.31) | 1.11 (0.79; 1.58) | 533 | Ref. | 1.49 (0.93; 2.38) | 0.78 (0.56; 1.11) | 308 | 0.01 (-0.22; 0.23) |
| **Daily contact family/friends**† | | | | | | | | | | |
| Yes | 1911 | Ref. | Ref. | Ref. | 1350 | Ref. | Ref. | Ref. | 1122 | Ref. |
| No | 553 | Ref. | 1.06 (0.82; 1.36) | 0.80 (0.60; 1.08) | 366 | Ref. | **1.33 (1.00; 2.14)** | **1.35 (1.00; 1.81)** | 160 | -0.07 (-0.26; 0.13) |
| **Feeling lonely (1-5 scale)**‡ | 1694 | Ref. | 1.10 (0.97; 1.26) | 0.95 (0.81; 1.12) | 1717 | Ref. | 1.09 (0.94; 1.26) | **0.86 (0.75; 0.98)** | | **0.11 (0.04; 0.18)** |
| **Housing conditions** | | | | | | | | | | |
| **Lack of outdoor views** | | | | | | | | | | |
| No | 2655 | Ref. | Ref. | Ref. | 1914 | Ref. | Ref | Ref. | 1182 | Ref. |
| Yes | 212 | Ref. | 1.16 (0.81; 1.67) | 1.15 (0.77; 1.70) | 181 | Ref. | **1.50 (1.00; 2.34)** | 1.18 (0.82; 1.71) | 103 | -0.10 (-0.34; 0.13) |
| **Lack of terrace/balcony** | | | | | | | | | | |
| No | 2020 | Ref. | Ref. | Ref. | 1535 | Ref. | Ref. | Ref. | 959 | Ref. |
| Yes | 847 | Ref. | 1.10 (0.89; 1.36) | 1.22 (0.97; 1.52) | 560 | Ref. | 1.10 (0.81; 1.49) | 1.09 (0.87; 1.38) | 326 | 0.01 (-0.14; 0.15) |
| **Lack of garden/yard** | | | | | | | | | | |
| No | 834 | Ref. | Ref. | Ref. | 398 | Ref. | Ref. | Ref. | 173 | Ref. |
| Yes | 2033 | Ref. | 0.59 (0.47; 0.74) | 1.86 (0.67; 1.09) | 1697 | Ref. | 0.86 (0.61; 1.25) | 0.82 (0.64; 1.07) | 1112 | -0.10 (-0.34; 0.13) |
| **Internet access** | | | | | | | | | | |
| Yes | 1865 | Ref. | Ref. | Ref. | 1499 | Ref. | Ref. | Ref. | 1026 | Ref. |
| No | 1002 | Ref. | 1.37 (1.09; 1.72) | 1.22 (0.96; 1.56) | 596 | Ref. | 1.10 (0.79; 1.54) | 0.86 (0.66; 1.12) | 259 | 0.05 (-0.11; 0.22) |

| | Changes in night-time sleep | | | | Changes in sleep quality | | | | | |
|---|---|---|---|---|---|---|---|---|---|---|
| | n | No changes | Worsening # | Improvement # | n | No changes | Worsening ## | Improvements ## | n | Changes in nº of poor sleep indicators### |
| | | | OR (95%CI) | OR (95%CI) | | | OR (95%CI) | OR (95%CI) | | $\bar{x}$ (95%CI) |
| | | n=971 | n=2020 | n=802 | | n=1180 | n=282 | n=633 | | n=1285 |
| **Too much noise** | | | | | | | | | | |
| No | 2778 | Ref. | Ref. | Ref. | 2034 | Ref. | Ref. | Ref. | 1254 | Ref. |
| Yes | 89 | Ref. | 1.03 (0.60; 1.76) | 0.75 (0.41; 1.40) | 61 | Ref. | **1.99 (1.00; 4.07)** | 1.56 (0.86; 2.85) | 31 | **0.47 (0.05; 0.89)** |
| **Lifestyle-behaviours** | | | | | | | | | | |
| **Smoking** | | | | | | | | | | |
| Never smokers | 1726 | Ref. | Ref. | Ref. | 1193 | Ref. | Ref. | Ref. | 660 | Ref. |
| Former smokers | 242 | Ref. | 0.95 (0.75; 1.22) | 1.03 (0.79; 1.35) | 700 | Ref. | 0.97 (0.69; 1.36) | 1.18 (0.92; 1.51) | 509 | 0.02 (-0.13; 0.16) |
| Smoker | 881 | Ref. | 0.95 (0.65; 1.39) | **1.73 (1.19; 2.50)** | 193 | Ref. | 0.77 (0.45; 1.33) | 1.07 (0.73; 1.55) | 116 | -0.12 (-0.35; 0.11) |
| **Alcohol intake** | | | | | | | | | | |
| Non drinker | 1231 | Ref | Ref. | Ref. | 778 | Ref. | Ref. | Ref. | 542 | Ref. |
| Drinker, not daily | 566 | Ref | 0.83 (0.63; 1.10) | 1.02 (0.75; 1.38) | 485 | Ref. | 1.16 (0.79; 1.70) | **1.47 (1.10; 1.94)** | 360 | 0.07 (-0.08; 0.23) |
| Drinker, daily | 671 | Ref | 0.82 (0.63; 1.08) | 0.97 (0.72; 1.31) | 456 | Ref. | 1.46 (0.97; 2.21) | 1.17 (0.86.1.59) | 383 | 0.14 (-0.03; 0.30) |
| **MEDAS** | | Ref | 1.03 (0.97; 1.09) | 0.98 (0.92; 1.04) | 2095 | Ref. | 0.94 (0.87; 1.02) | **1.10 (1.03; 1.16)** | 1285 | 0.00 (-0.15; 0.15) |
| **PA (cohort-specific quartiles)** | | | | | | | | | | |
| 1st quartile (less) | 725 | Ref. | Ref. | Ref. | 531 | Ref. | Ref. | Ref. | 334 | Ref. |
| 2nd quartile | 740 | Ref. | 0.77 (0.59; 1.02) | 0.89 (0.67; 1.18) | 541 | Ref. | 1.24 (0.83; 1.86) | 0.85 (0.65; 1.13) | 331 | 0.11 (-0.06; 0.29) |
| 3rd quartile | 743 | Ref. | 0.77 (0.58; 1.01) | 0.81 (0.60; 1.09) | 550 | Ref. | **1.62 (1.09; 2.42)** | 0.86 (0.64; 1.15) | 332 | **0.22 (0.03; 0.41)** |
| 4th quartile | 659 | Ref. | 0.86 (0.64; 1.14) | 0.90 (0.66; 1.22) | 473 | Ref. | 1.45 (0.94; 2.21) * | 0.82 (0.61; 1.11) | 288 | **0.30 (0.10; 0.49)** |
| **BMI** | | | | | | | | | | |
| Normoweight | 649 | Ref | Ref. | Ref. | 551 | Ref. | Ref. | Ref. | 356 | Ref. |
| Overweight | 1341 | Ref | 0.95 (0.74; 1.22) | 1.13 (0.86; 1.49) | 992 | Ref. | 0.84 (0.60; 1.19) | 0.91 (0.71; 1.16) | 635 | 0.05 (-0.11; 0.20) |
| Obese | 877 | Ref | 0.86 (0.65; 1.13) | 1.12 (0.83; 1.52) | 552 | Ref. | 0.84 (0.57; 1.23) | **0.74 (0.55; 1.00)** | 294 | 0.07 (-0.12; 0.26) |
| **ST (cohort-specific quartiles)**§ | | | | | | | | | | |
| 1st quartile | 610 | Ref. | Ref. | Ref. | 613 | Ref. | Ref. | Ref. | 338 | Ref. |
| 2nd quartile | 490 | Ref. | 0.80 (0.58; 1.10) | 0.93 (0.65; 1.32) | 480 | Ref. | 1.25 (0.86; 1.80) | 1.13 (0.85; 1.51) | 307 | 0.08 (-0.10; 0.26) |
| 3rd quartile | 528 | Ref. | 0.77 (0.56; 1.06) | 0.92 (0.66; 1.29) | 523 | Ref. | 0.94 (0.64; 1.38) | **1.41 (1.07; 1.86)** | 325 | 0.03 (-0.15; 0.21) |
| 4th quartile | 476 | Ref. | 0.95 (0.69; 1.30) | 0.86 (0.60; 1.24) | 479 | Ref. | 1.04 (0.71; 1.54) | 1.08 (0.81; 1.45) | 315 | 0.02 (-0.17; 0.20) |
| **Sleep characteristics** | | | | | | | | | | |
| **Night-time sleep** | | | | | | | | | | |
| Normal sleep | 2140 | - | ; | ; | 1614 | Ref. | Ref. | Ref. | 1024 | Ref. |
| Short sleep (≤6 h) | 488 | - | ; | ; | 351 | Ref. | **1.51 (1.08; 2.13)** | 1.30 (0.98; 1.71) | 206 | **0.39 (0.20, 0.58)** |
| Long sleep (≥9 h) | 239 | - | ; | ; | 130 | Ref. | 0.70 (0.37; 1.35) | 0.99 (0.65; 1.49) | 55 | -0.12 (-0.44; 0.19) |
| **Day-time sleep**‖ | | | | | | | | | | |
| No | 582 | Ref. | Ref. | Ref. | 573 | Ref. | Ref. | Ref. | 388 | Ref. |
| Short nap | 673 | Ref. | 0.78 (0.56; 1.08) | 1.04 (0.74; 1.48) | 669 | Ref. | 0.98 (0.68; 1.40) | 0.83 (0.63; 1.10) | 610 | 0.07 (-0.07; 0.22) |
| Long nap | 339 | Ref. | 1.12 (0.78; 1.61) | 1.06 (0.72; 1.57) | 89 | Ref. | 0.92 (0.60; 1.43) | 0.78 (0.57; 1.11) | 217 | 0.05 (-0.14; 0.26) |
| Very long nap | 94 | Ref. | 1.18 (0.64; 2.17) | 1.71 (0.94; 3.12) | 27 | Ref. | 0.86 (0.39; 1.88) | 0.77 (0.46; 1.31) | 67 | -0.04 (-0.35; 0.27) |

| | Changes in night-time sleep | | | | | | Changes in sleep quality | | | | |
|---|---|---|---|---|---|---|---|---|---|---|---|
| | n | No changes | Worsening # | | Improvement # | n | No changes | Worsening ## | Improvements ## | n | Changes in nº of poor sleep indicators### |
| | | | OR (95%CI) | | OR (95%CI) | | | OR (95%CI) | OR (95%CI) | | $\bar{x}$ (95%CI) |
| | | n=971 | n=2020 | | n=802 | | n=1180 | n=282 | n=633 | | n=1285 |
| **Overall sleep quality§** | | | | | | | | | | | |
| Very good | 408 | Ref. | 0.99 (0.73; 1.33) | | 0.64 (0.44; 0.93) | 411 | Ref. | - | - | 140 | - |
| Good | 1210 | Ref. | Ref. | | Ref. | 1212 | Ref. | - | - | 834 | - |
| Poor/very poor | 57 | Ref. | **0.18 (0.05; 0.60)** | * | 0.88 (0.42; 1.85) | 61 | Ref. | - | - | 28 | - |
| **Overall health** | | | | | | | | | | | |
| 1$^{st}$ quartile | 544 | Ref. | Ref. | | Ref. | 594 | Ref. | Ref. | Ref. | | Ref. |
| 2$^{nd}$ quartile | 544 | Ref. | 0.85 (0.61; 1.17) | | 0.88 (0.63; 1.25) | 453 | Ref. | 0.86 (0.59; 1.27) | 0.91 (0.66; 1.25) | 309 | **-0.28 (-0.47; -0.08)** |
| 3$^{rd}$ quartile | 597 | Ref. | 0.89 (0.65; 1.23) | | 0.89 (0.63; 1.26) | 503 | Ref. | 0.73 (0.48; 1.11) | 0.80 (0.57; 1.11) | 329 | **-0.44 (-0.64; -0.23)** |
| 4$^{th}$ quartile (best) | 1182 | Ref. | **0.73 (0.53; 1.00)** | * | 0.82 (0.58; 1.16) | 545 | Ref. | 0.70 (0.46; 1.08) | 1.15 (0.83; 1.59) | 340 | **-0.40 (-0.62; -0.18)** |
| **Pain scale¶** | 1025 | Ref. | 1.01 (0.92; 1.11) | | 1.01 (0.92; 1.12) | 1029 | Ref. | **1.11 (1.01; 1.23)** | 1.05 (0.97; 1.15) | 307 | **0.06 (0.03; 0.10)** |
| **Chronic morbidities** | | | | | | | | | | | |
| Diabetes | 490 | Ref. | 0.92 (0.71; 1.20) | | 1.09 (0.83; 1.43) | 346 | Ref. | 1.33 (0.94; 1.89) | 0.97 (0.72; 1.29) | 230 | 0.07 (-0.10; 0.24) |
| Hypertension | 1582 | Ref. | 1.00 (0.82; 1.23) | | 1.12 (0.90; 1.39) | 1046 | Ref. | **1.47 (1.10; 1.96)** | 1.11 (0.90; 1.38) | 699 | **0.15 (0.02-0.28)** |
| CVD† | 178 | Ref. | 0.67 (0.44; 1.02) | | 0.76 (0.49; 1.18) | 102 | Ref. | 1.18 (0.64; 2.17) | 0.93 (0.56; 1.53) | 53 | -0.03 (-0.35; 0.29) |
| Cancer† | 173 | Ref. | 1.27 (0.86; 1.88) | | 1.20 (0.78; 1.83) | 112 | Ref. | 1.18 (0.66; 2.14) | 1.30 (0.83; 2.04) | 71 | -0.05 (-0.33; 0.23) |
| Osteomuscular disease | 1089 | Ref. | 0.89 (0.71; 1.10) | | 0.97 (0.77; 1.23) | 895 | Ref. | 0.87 (0.65; 1.17) | 0.87 (0.69; 1.09) | 620 | **0.18 (0.04; 0.32)** |
| Depression | 342 | Ref. | 0.99 (0.72; 1.36) | | 1.05 (0.75; 1.46) | 243 | Ref. | **1.58 (1.05; 2.39)** | **1.52 (1.07; 2.15)** | 155 | **0.25 (0.03; 0.47)** |
| **Mobility limitations¶** | 459 | Ref. | 0.65 (0.63; 0.98) | | 1.15 (0.74; 1.77) | 458 | Ref. | 1.19 (0.96; 1.85) | 1.21 (0.84; 1.74) | 458 | -0.08 (-0.25; 0.08) |
| **Negative ageing experience scale** | 1284 | Ref. | 0.99 (0.84; 1.16) | | 1.05 (0.88; 1.27) | 1287 | Ref. | 1.11 (0.93; 1.34) | 0.93 (0.80; 1.08) | 1287 | 0.05 (-0.02; 0.12) |
| **Cantril ladder‡** | 1691 | Ref. | 1.03 (0.96; 1.11) | | 1.01 (0.93; 1.10) | 1710 | Ref. | 0.99 (0.91; 1.08) | **1.11 (1.03; 1.19)** | 1287 | -0.03 (-0.06; 0.01) |
| **Low MMSE scores (<23)‡** | 63 | Ref. | 0.90 (0.47; 1.73) | | 0.79 (0.33; 1.88) | 56 | Ref. | 1.04 (0.45; 2.41) | 1.27 (0.66; 2.42) | 34 | 0.35 (-0.06; 0.75) |
| **Years since baseline exam** | 2867 | Ref. | 0.86 (0.70; 1.05) | | 1.04 (0.93; 1.15) | 2095 | Ref. | 0.81 (0.58; 1.13) | 0.97 (0.87; 1.07) | 1285 | 0.02 (-0.05; 0.08) |
| **Week state of alarm** | 2867 | Ref. | 0.97 (0.86; 1.08) | | 0.88 (0.72; 1.09) | 2095 | Ref. | 0.99 (0.86; 1.13) | 0.84 (0.65; 1.09) | 1285 | 0.00 (-0.15; 1.92) |

OR: Odds Ratio; 95%CI: 95% Confidence interval; $\bar{x}$: Mean change. Confidence intervals are represented by the lower limit and the upper limit with either a hyphen (when the main estimates is an OR) or a semicolon (when the main estimate is a difference in means)

MEDAS: Mediterranean Diet Assessment Score; BMI: Body Mass Index; PA: Physical activity; ST: Sedentary time; MMSE: Mini Mental State Examination.

Note: Figures highlighted in bold show a statistically significant association.

Note: Figures highlighted in bold show a statistically significant association.

**#**Worsening was defined by having a healthy night-time time sleep duration (6-9 hours) in the pre-confinement period but not during confinement. Improvements were defined by sleeping less than 6 hours or more than 9 in the pre-confinement period and 6-9 during confinement

**##**Worsening was defined by experiencing increases in the "poor sleep quality score", and improvements as experiencing decreases in the same score.

**###**This score ranged from 0 to 7. Those who answered "sometimes" or "almost always" to the items "difficulty falling asleep", "awakening during night-time", "early awakening with difficulty getting back to sleep" "use of sleeping pills", "being so sleepy at daytime as to need a nap" or "not feeling rested in the morning", as well as those with an Epworth Sleepiness Scale score >10, received 1 point; their counterparts received 0 points. This score was only available in *Seniors ENRICA-2* and *Exernet*

†Information not available in EXERNET; ‡Information only available in *Seniors ENRICA-2* and *Edad con Salud*; §Information not available in *TSHA*; ‖Information only available in *Seniors ENRICA-2* and *Exernet* ¶Information only available in *Seniors-ENRICA-2.*

All models were adjusted for baseline age, sex (men or women), educational level (primary or less, secondary, or university), civil status (married, widowed, never married, divorced), housing conditions, smoking status (never, former, or current), alcohol drinking (never, former, moderate, or heavy), adherence to the Mediterranean diet (MEDAS score), body mass index (normoweight, overweight, or obese), physical activity (quartiles), hours of sleep at night (normal, short sleep, long sleep), chronic comorbidities (hypertension, diabetes, cardiovascular disease, cancer, osteo-muscular disease and depression), time since last follow-up visit, cohort of study and week of the state of alarm.

*p-value for trend <0.05
